# Early underdetected dissemination across countries followed by extensive local transmission propelled the 2022 mpox epidemic

**DOI:** 10.1101/2023.07.27.23293266

**Authors:** Miguel I. Paredes, Nashwa Ahmed, Marlin Figgins, Vittoria Colizza, Philippe Lemey, John T. McCrone, Nicola Müller, Cécile Tran-Kiem, Trevor Bedford

## Abstract

The World Health Organization declared mpox a public health emergency of international concern in July 2022. To investigate global mpox transmission and population-level changes associated with controlling spread, we built phylogeographic and phylodynamic models to analyze MPXV genomes from five global regions together with air traffic and epidemiological data. Our models reveal community transmission prior to detection, changes in case-reporting throughout the epidemic, and a large degree of transmission heterogeneity. We find that viral introductions played a limited role in prolonging spread after initial dissemination, suggesting that travel bans would have had only a minor impact. We find that mpox transmission in North America began declining before more than 10% of high-risk individuals in the USA had vaccine-induced immunity. Our findings highlight the importance of broader routine specimen screening surveillance for emerging infectious diseases and of joint integration of genomic and epidemiological information for early outbreak control.

## Introduction

Mpox is a viral zoonotic disease caused by the mpox virus (MPXV), previously referred to as monkeypox virus, that is endemic to West and Central Africa (1,2). Prior to 2022, most cases of mpox outside of endemic regions occurred in individuals with either a recent travel history to Nigeria or with an exposure to live animals from endemic areas. On May 7, 2022, an individual with a travel history to Nigeria was diagnosed with mpox in the United Kingdom (UK) (3). Following this initial detection, the number of mpox cases without a travel history to endemic countries began to increase rapidly in various regions around the globe consistent with epidemic human-to-human spread (3). As of July 19, 2023, the Centers for Disease Control and Prevention (CDC) reported 88,549 cases of mpox worldwide since Jan 2022 (4).

The 2022 mpox epidemic was characterized by human-to-human spread outside of endemic areas, mostly in men who have sex with men (MSM), that resulted in a less severe illness presentation compared to what was seen in historical short human-to-human transmission chains following repeated zoonoses (2,3,5). The long incubation period of 5-21 days (3,6), as well as the atypical and less severe illness presentation suggests that mpox may have spread undetected prior to initial case discovery. Presymptomatic transmission of mpox has also been documented, suggesting that the epidemic was at least partially fueled by transmission occurring prior to symptom onset (7–9).

The WHO declared mpox to be a public health emergency of international concern on July 23, 2022, promoting investigations into disease spread, the use of vaccines to control transmission, and potential guidelines for international travel (10). Individual countries began vaccination efforts in an attempt to curb mpox spread but have been criticized for long delays in starting effective vaccination campaigns in high-risk areas (11). To date, it is still unclear to what extent continued international travel contributed to the explosive spread of mpox in various global regions and whether or not national vaccination campaigns were wholly responsible for controlling the epidemic.

Genomic epidemiology is uniquely poised to explore global and regional transmission dynamics through the joint integration of viral genomic information and epidemiological metadata. This approach augments traditional public health surveillance, especially when case-based surveillance is limited (12). While a few studies have looked into the regional spread of mpox at various stages of the 2022 epidemic (13–16), most relied on very few pathogen genomes. Overall, the extent of undetected mpox spread and the effectiveness of proposed interventions have yet to be examined. Here we employ recent advances in phylogeographic and phylodynamic methods to estimate changes in case detection rate, the impact of underdetection on transmission, and the role of introductions in promoting local community spread in various global regions. We also examine the impact of vaccination campaigns on epidemic growth and decay in North America as well as estimate the degree of transmission heterogeneity in the declining phase of the epidemic.

## Results

### Early mpox spread in Western Europe sparks prolonged outbreaks in Southern Europe, North America and South America

Following initial detection in the UK on May 7, 2022, the number of mpox cases reported worldwide grew rapidly (Figure 1). In early May, reported cases were found mainly in Western and Southern, and then Central, Europe where the epidemic peaked around mid-July (Figures 1A, B). Beginning in mid-May, however, cases began to be reported in North America, which ultimately led to the largest number of reported cases of any global region studied, peaking at the beginning of August. Around the same time as the North American peak, cases were detected and started rising in South America, which substantially contributed to the later tail of the 2022 mpox epidemic. Similarly, the number of sequences collected increased as more cases were detected, with heterogeneity between regions and North America (primarily the US) submitting the largest number of sequences to GenBank. (Figures 1C, D).

**Figure 1.**
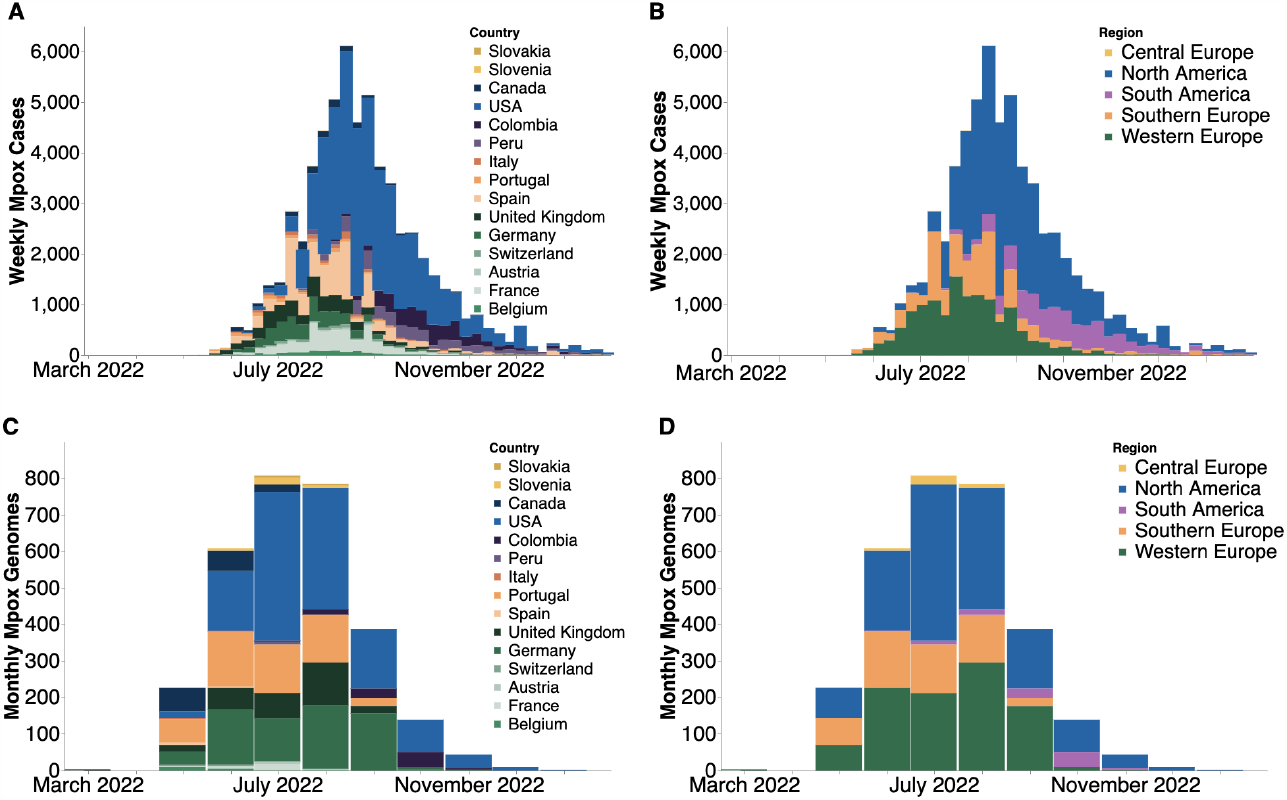
Case counts and publicly available sequences by geographic region. (**A, B**) Confirmed positive weekly mpox cases by country (**A**) and global region (**B**) smoothed using a 7 day rolling average on daily data and then aggregating into weekly counts. Only countries with greater than 5 sequences on GenBank were included. (**C, D**) Monthly count of publicly-available MPXV genomes found on GenBank by country (**C**) and global region (**D**).

To investigate the spread of mpox throughout the course of the epidemic across global regions, we employed a phylogeographic approach with an asymmetrical discrete trait model on 1004 publicly available MPXV sequences subsampled based on confirmed case counts (Figure 2, Figure S1A) in order to infer the global region of origin for all internal ancestral nodes. We chose a case count weighted subsampling scheme since discrete trait analysis assumes that sample sizes across subpopulations are proportional to their relative population prevalence (17). Due to the low number of recorded cases in Central Europe, no sequences from that region were included in the final subset (Figure S1B). We infer that the most recent common ancestor (MRCA) of the epidemic existed between March 9th and March 27th, 2022 (95% HPD) and phylogeographic estimation assigns this ancestor to Western Europe. We infer the evolutionary clock rate to be 8. 41× 10^−5^ (95% HPD 7. 71× 10^−5^ to 9. 10× 10^−5^) substitutions per site per year or approximately 16.8 substitutions per genome per year. Alternative phylodynamic models place the MRCA in November or December 2021, but show similar estimates of clock rate (Table S2).

**Figure 2.**
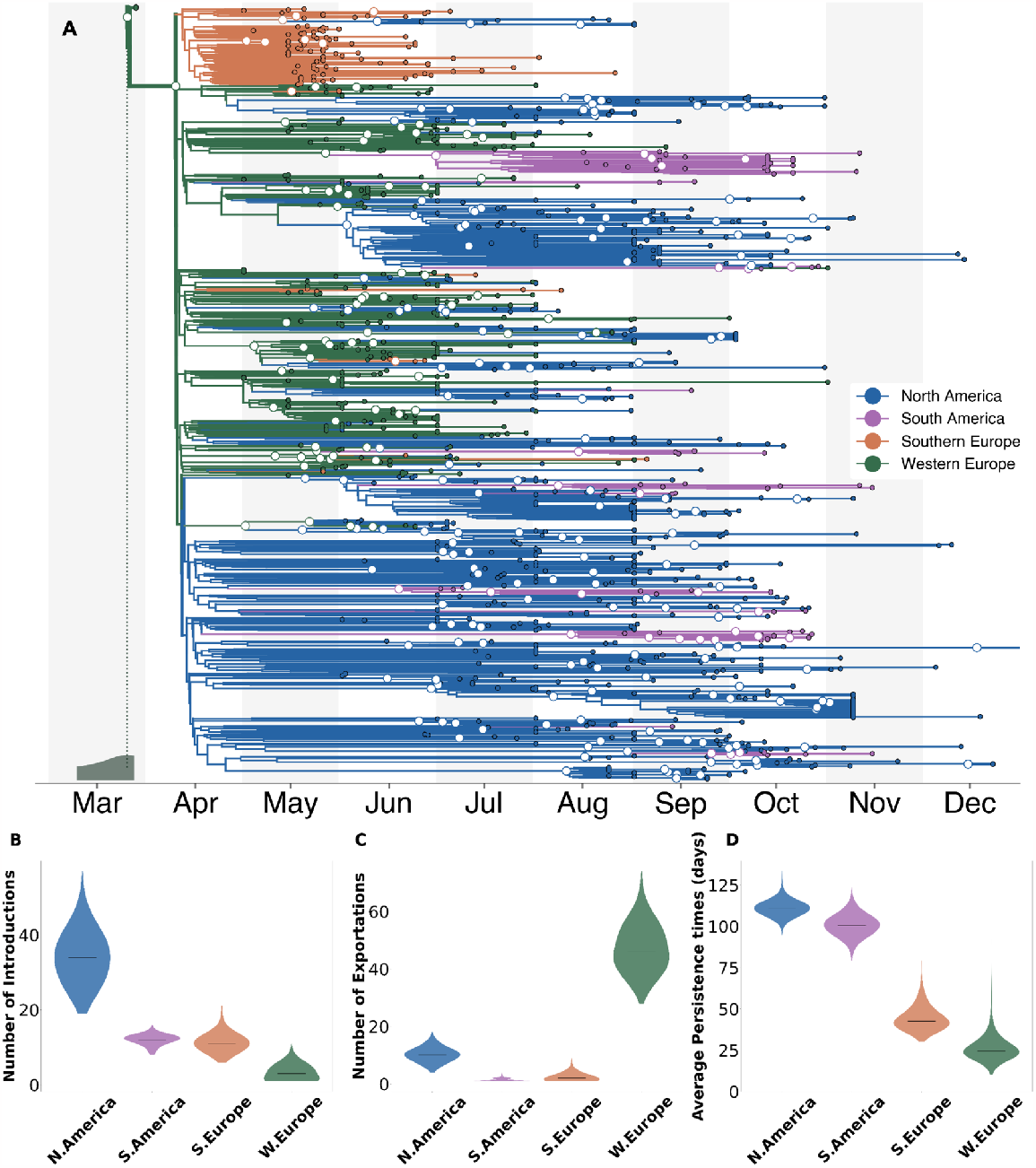
Phylogeographical estimates of MPXV spread in 4 global regions. (**A**) The maximum clade credibility tree summary of the Bayesian inference conducted using asymmetric discrete trait analysis and Skygrid prior on 1004 sequences. Colors correspond to the regions in the legend. Ancestral nodes with greater than 50% posterior support are highlighted with a white circle overlaid. Inset histogram on bottom left corner shows 95% interval for the time to most recent common ancestor (TMRCA)(**B-D**) Estimated number of introductions (**B**), exports (**C**), and average time of local persistence in days (**D**) for each global region. Horizontal black line denotes median estimates.

We observe strong population structure where single introductions often result in large local clades. These large local clades suggest that local spread played a considerable role in their respective regional outbreaks. We find rapid early spread in Western Europe lead to a high number of introductions to other global regions (46 introduction events, IQR: 41-53), seeding regional outbreaks (Figure 2C). Our findings also show evidence of repeated dissemination into North America and subsequent sustained community transmission as North America had the highest median number of viral importations and longest median viral persistence time (111 days, IQR: 108-114) (Figure 2B,D).

To test the appropriateness and accuracy of the phylogeographic inference, we repeated the analysis in which 10% (100 tips in total) of the sequence locations were masked. We then inferred these locations via the same phylogeographic approach and found that the model correctly inferred 93% of the masked tip locations, suggesting a strong genomic signal (Figure S2). Additionally, we also repeated our analysis using an equal spatiotemporal subsampling scheme for every year-week in the studied time period as well as by subsampling directly from each region rather than from individual countries and found highly similar estimates of the MRCA and of patterns of introductions, exportations, and persistence regardless of the subsampling scheme used (Figure S3)

### Rapid early spread characterized by significant underdetection of cases

In order to analyze within-region transmission dynamics, improve robustness to sampling bias, and enhance inference via the joint integration of genomic and epidemiological metadata, we then employed an approximate structured coalescent (MASCOT) with a generalized linear model (GLM) approach with estimated prevalence and air passenger data as empirical predictors on 587 sequences in order to infer the effective population size and migration rates within and between each region, respectively (Figure S4A). We also included a predictor for each month within the time period studied to account for potential changes in case detection over time. The included sequences were subsampled with equal temporal weighting to increase representation of undersampled regions such as Central Europe (Figure S1A, C, see STAR Methods for more information). Despite the improved computational efficiency of MASCOT over standard structured coalescent approaches (18), parameter inference under MASCOT-GLM is still computationally demanding compared to discrete trait analysis. For reference, the runtime for our main DTA analysis with 1004 sequences was about 12.26 hours/million states while for MASCOT-GLM with only 587 sequences it was about 16.45 hours/million states. Using a minimum of 5 * 10^7^ MCMC steps to promote convergence, these runtimes translate to 25.5 days of computational demand for our main DTA analysis and 34.3 days for our main MASCOT-GLM analysis. As such, we reduced the number of sequences to 587 for MASCOT-GLM to allow for inference within actionable timescales (Figure S1C). Additionally, the MASCOT-GLM subsampling scheme is different from the subsampling for the DTA analysis as the structured coalescent is more robust to differences in sampling across regions and is subsequently informed by regional prevalence (17,19). We used a GLM approach in order to draw inferential power from relevant predictors and reduce uncertainty relative to inferences using the coalescent alone. After separating out each introduction and its inferred descendents from the maximum clade credibility tree and comparing them to confirmed case counts, we see strong evidence of viral circulation before initial detection in each global region (Figure 3A). Additionally, we revealed that the largest downstream outbreak clusters arise from introductions prior to detection from public health surveillance while introductions after detection are more likely to be a single case and extinguish quickly (Figure S4B).

**Figure 3.**
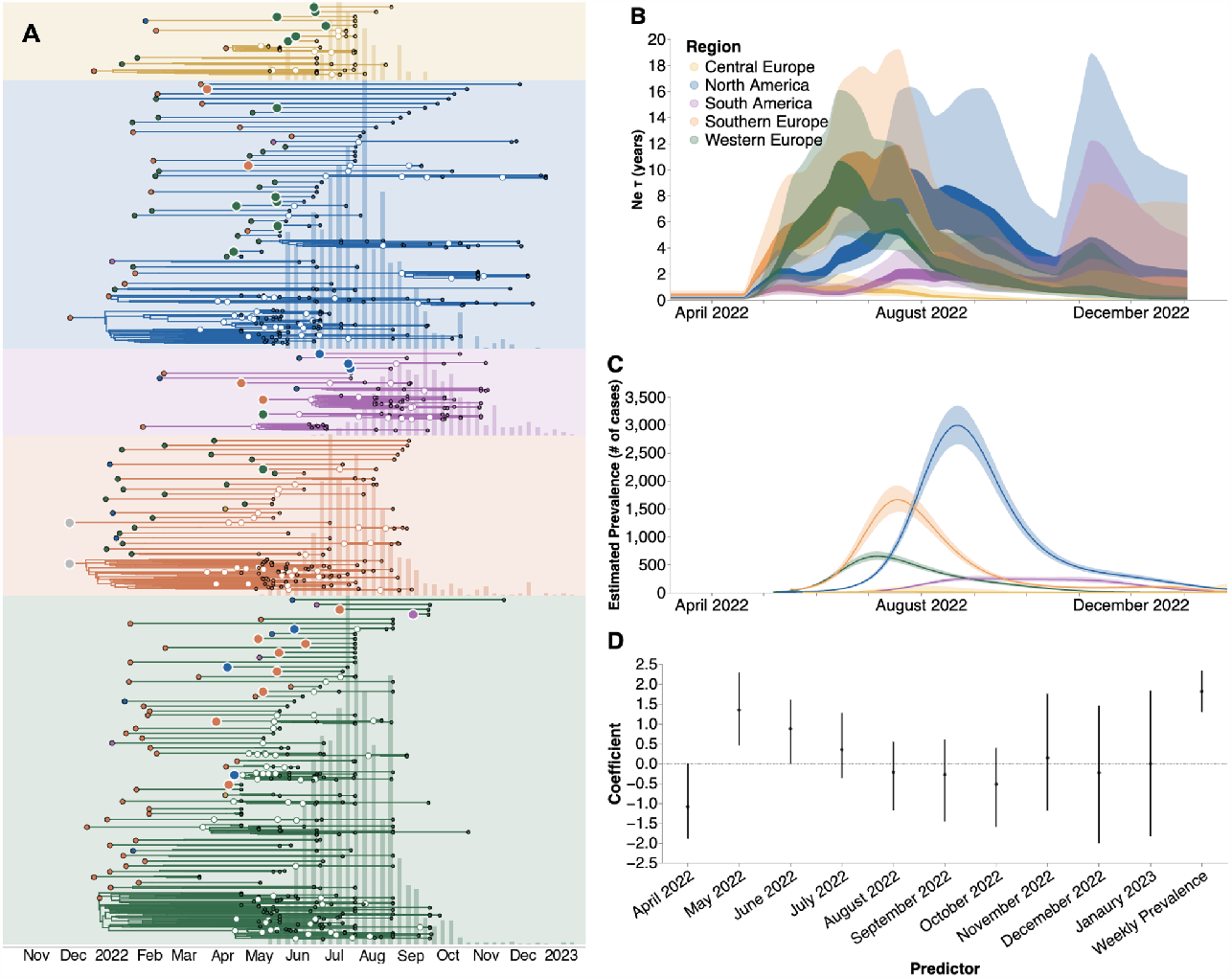
Phylodynamic investigation reveals underdetection of mpox. (**A**) Regional specific introductions and the resulting outbreak clusters extracted from the maximum clade credibility tree summary of the Bayesian inference conducted using MASCOT-GLM on 587 sequences. Colors correspond to the regions in the legend. Ancestral nodes with greater than 50% posterior support are highlighted with a white circle overlaid. (**B**) Estimates of effective population sizes (*Neτ* in years) from April 2022 through December 2024 using 550 sequences subsampled equally throughout time. The coalescent time scale depends on both effective population size *Ne* (number of effective individuals) and on generation time τ (years per generation), resulting in Neτ being a measure of coalescent time scale in years (20). (**C**) Regional prevalence (in number of cases, interpreted as census population size N) estimated independently using publicly-available case counts, and (**D**) Estimates of model predictor coefficients for *Ne* estimation. All of the predictors displayed on the x-axis were included in the analytic model. Dark line represents median estimates, light bands represent 95% HPD.

We sought to investigate the extent of underdetection in each region by comparing the MASCOT-GLM estimates of effective population size *Ne* (Figure 3B) with the prevalence estimated solely from case counts (Figure 3C), which we assume to approximate the census population size. Of note, the MASCOT-GLM estimates are informed by prevalence as an empirical predictor, allowing us to assume that differences between the coalescent-derived *Ne* and case-based prevalence estimates could be due to differential case reporting. While both estimates show regional peaks at similar points in time, we find a divergence between the two estimates in the early months of the outbreak – May, June, July 2022 – where our coalescent-derived *Ne* show continuous viral epidemic growth before case-based prevalence counts report any cases detected by local public health authorities, suggesting significant underdetection of cases in these months. This observation is supported by the estimated coefficients of the monthly predictors that show the direction and magnitude of each predictor’s effect on the inference of regional *Ne*. Figure 3D shows that the predictors for the months of May, June, and July 2022 had a strong positive effect on predicting regional *Ne*. By August 2022, however, when a substantial number of cases had been detected in all five regions, we see that our model no longer finds the monthly predictors to be required, implying that prevalence estimates are sufficient to describe *Ne*. The strong positive effect of the monthly predictors from May through July, even in the presence of competing information from the prevalence predictor, suggests significant underreporting of cases in these first few months. For comparison with our predictor-informed MASCOT-GLM model, a strictly coalescent-based model without predictors (Figure S5) shows similar trends in *Ne* but displays a larger degree of uncertainty, supporting the use of empirical predictors to inform our inference.

### After initial dissemination, viral importations had limited impact on local spread

When analyzing transmission chains resulting from introductions (Figure 3A), we identified a bimodal pattern in each region, where most viral introductions resulted in a single imported case while a very small number of introductions spark explosive and widespread local transmission. Upon identifying the regional introductions with the highest posterior support in our MCC tree, we find that introductions that occurred early in the global outbreak lead to larger and more persistent transmission chains, while those introductions that occurred after initial public health detection in each region resulted in smaller outbreaks that extinguished faster (Figure 4A, Figure S4B). The clear negative correlation between time of introduction and persistence of downstream transmission chains remains even without the influence of the two large transmission chains following the first two inferred introductions (Figure S4C). We also found that air passenger volumes between each global region were a significant positive predictor of viral migration between each region, highlighting the importance of regional connectivity in promoting international viral spread (Figure 4B).

**Figure 4.**
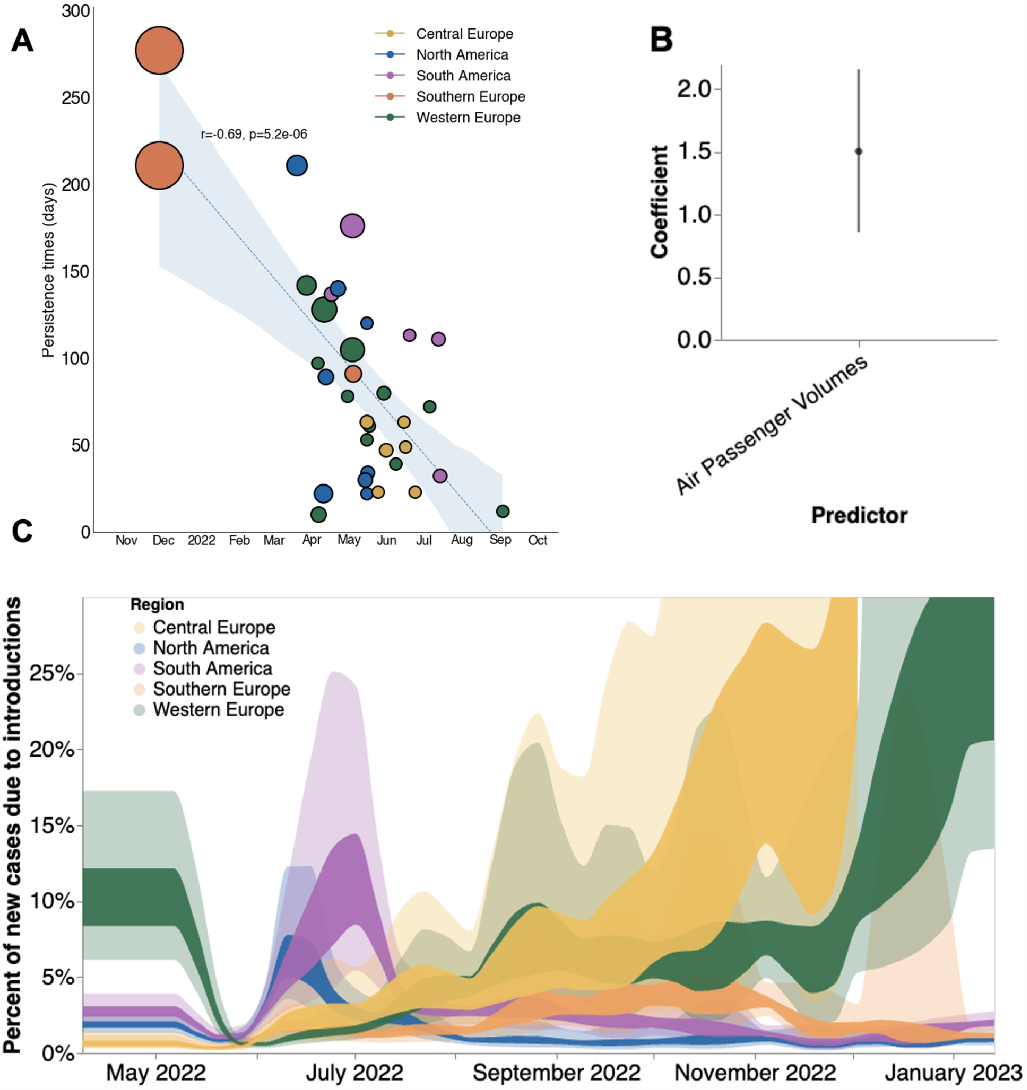
Phylodynamic estimates of MPXV transmission dynamics in 4 global regions. (**A**) Relationship between estimated date of introduction and persistence time. Each circle represents a single viral introduction with greater than 50% posterior support into the region denoted by the color (i.e. a green point represents an introduction into Western Europe). The size of each point is proportional to the size of the outbreak cluster resulting from each introduction with larger circles representing more resulting downstream tips. Blue dashed line represents the linear best fit line using Pearson’s correlation. Blue shaded region denotes the variability of the line and the resulting estimates from Pearson’s correlation are shown in text above the shaded region. (**B)** Estimates of model predictor coefficients for migration rate estimation. Error bars denote 95% HPD interval for the magnitude of predictor coefficient (**C**) Percentages of new cases due to introductions were estimated as the relative contribution of introductions to the overall number of infections in the region. The inner area denotes the 50% HPD interval and the outer area denotes the 95% HPD interval. Estimates were smoothed using a 14 day rolling average.

We sought to estimate the relative contribution of introductions versus local community spread in driving the epidemic in each global region via inferred parameters from MASCOT-GLM. To quantify the impact of those introductions, we calculated the percentage of new cases from introductions in each region using the estimated changes in *Ne* over time, the rate of viral migration between regions, and the incubation and infectious periods distributions for mpox. We found that introductions played a relatively small role in each regional epidemic, with introductions resulting in an average of 1.5-10% of new cases over the time period studied (Figure 4C). We see the percentage of new cases due to viral introductions in North and South America peaking at the start of their respective epidemics and then quickly drop down once the epidemic begins to peak. This finding suggests that following the initial viral seeding from importation events, local transmission dominates and that viral introductions play a very limited role in the later stages of regional epidemics. We also see large variability in the contribution of introductions on local spread during the later months which could be driven by lack of genomic and case-based information at those time periods (Figure 4C).

To better understand transmission dynamics locally within each region, we computed *Rt*, the time-varying effective reproductive number, using the estimated growth rate derived from changes in effective population size (Figure 5). We also employed our estimates of the percentage of new cases that are due to introductions to calculate *Rt* without the influence of introductions (Figure S6A). Initially, we observe high *Rt* with viral establishment in each respective region followed by a subsequent rapid decrease in which most regions achieve *Rt* < 1 (signaling an declining epidemic) by September 2022. Initial Rt of 1.5–3 corresponds to an epidemic doubling every 5.6–22.1 days. Removing the contribution of introductions, however, has a very small effect on regional *Rt*, showing the limited impact of introductions on local viral spread after initial regional establishment (Figure S6A).

**Figure 5:**
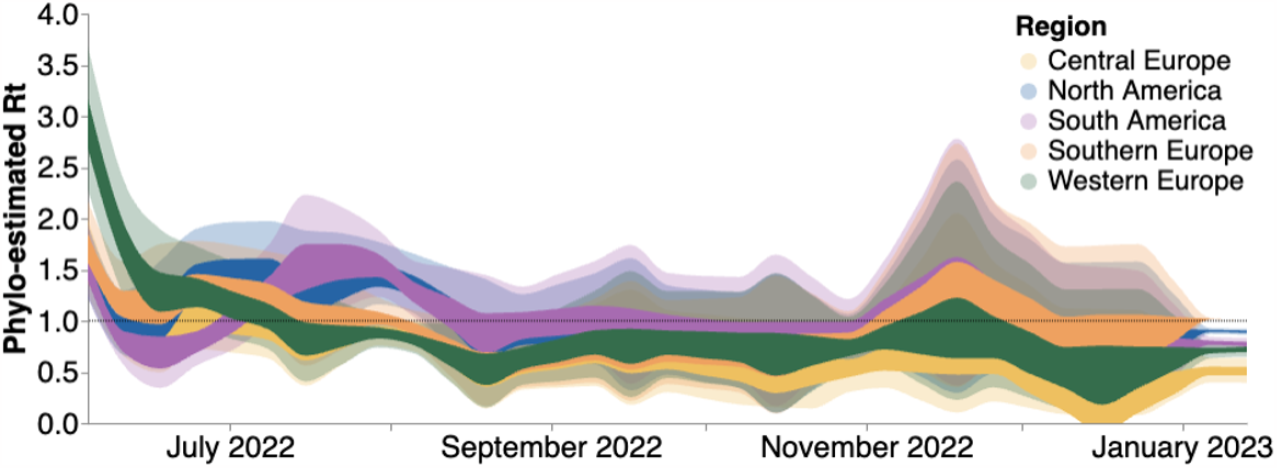
Estimates of time-varying reproductive number (*Rt*) in five global regions. Estimates of *Rt* from April 2022 through December 2023 via MASCOT-GLM using 587 sequences subsampled equally throughout time. The inner area denotes the 50% HPD interval and the outer area denotes the 95% HPD interval. Dashed line highlights an *Rt* value of 1 above which denotes an exponentially growing viral epidemic. Estimates were smoothed using a 14 day rolling average.

Additionally, we calculated *Rt* using only case counts (See Methods) to highlight the impact of accounting for underreporting on *Rt* estimation (Figure S6B). Compared to our phylodynamic estimates that take into account underreporting in the first months of the epidemic, renewal based methods for *Rt* estimation from only case counts significantly overestimate the transmissibility of mpox for every region.

### US vaccine campaigns had limited impact on curbing the North American outbreak

Given that North America bore the highest burden of mpox cases throughout the epidemic, we focused on this region to explore the role introductions had on prolonging the local epidemic as well as the impact of mpox vaccination on *Rt*. We find that introductions accounted for only an average of about 5-15% of local spread. By focusing on the declining half of the North American epidemic (dates laters than June 15, 2022), we additionally found that preventing introductions following the initial seeding event would have caused the *Rt* to fall below one only less than a week earlier (Figure S6A), highlighting the relatively low importance of introductions.

When we overlaid North American *Rt* estimates alongside the cumulative percentage of high-risk individuals in the USA with mpox vaccine-derived immunity (for description of the data and definitions, see Methods, under *Data Sources*), we found that *Rt* began declining prior to initiation of vaccination in the US (Figure 6A). Vaccine-induced immunity was estimated via a two week lag since the date of vaccination. North American *Rt* estimates fell below one near the middle of August 2022, when the cumulative percentage of high-risk individuals with vaccine-derived immunity was less than 8%. Under an SIR model of infectious disease dynamics, vaccine-derived immunity impacts disease transmission by removing individuals from the susceptible population in a linear fashion. Before there was any mpox vaccine-derived immunity in the US, North American *Rt* peaked at 1.49. Assuming a linear decrease in *Rt* as cumulative vaccine-derived immunity increased, we would expect *Rt* to fall below 1 only after greater than 33% of the high-risk population of the US developed immunity against mpox (Figure 6B, dashed gray line). When we compare the actual decay of *Rt* in North America, we find that *Rt* falls below one before about 10% of the high risk population developed immunity (Figure 6B, blue scatter points and red spline), implying that vaccination is not primarily responsible for the drop of *Rt* below 1. The decay of *Rt* in North America before a substantial percentage of high-risk individuals developed vaccine-related immunity remains clear even when assuming for no lag or a one week lag after vaccination for the development of immunity (Figure S6C-D). Of note, we were only able to publicly access vaccination information for the US via the CDC but our regional *Rt* analysis for North America includes viral dynamics for both the US and Canada.

**Figure 6.**
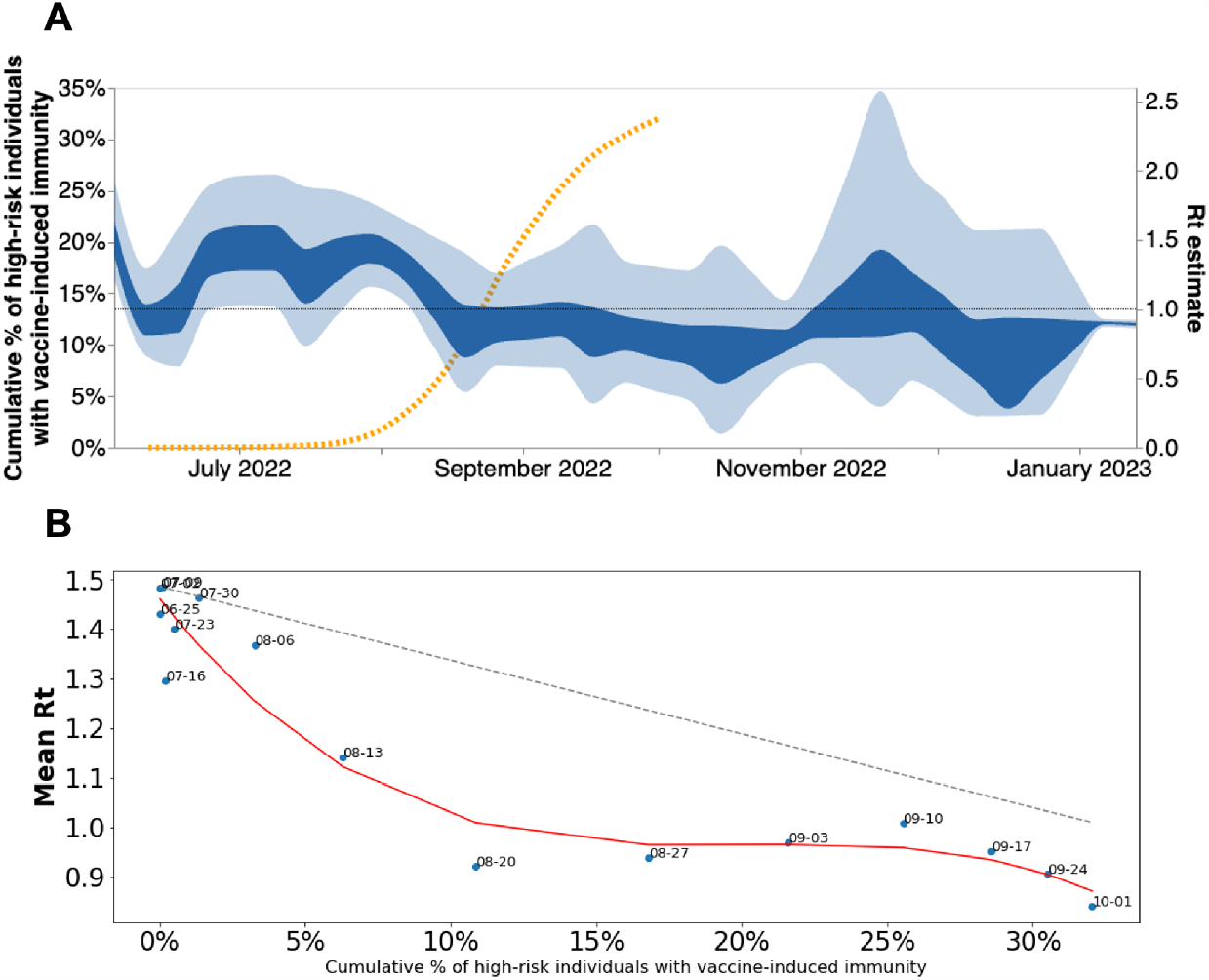
North American MPXV local transmission dynamics. (**A**) North American *Rt* estimated via phylodynamics (solid bands). Dashed orange line indicates the cumulative percentage of high-risk individuals with vaccine-induced immunity in the US. (**B**) Scatter plot comparing mean *Rt* calculated via MASCOT-GLM for North America vs cumulative percentage of high risk individuals with vaccine-induced immunity in the United States. Vaccine-induced immunity was estimated via a two week lag since the date of vaccination. Red line indicates the best fit spline for scattered points. Dashed gray line indicates expected linear decrease in *Rt* with increasing vaccine-immunity assuming SIR dynamics. Over each point are the dates that correspond to the mean *Rt* and percent of immunity at that moment.

### High degree of transmission heterogeneity observed in the declining phase of the mpox epidemic

Upon separating out each introduction and its inferred descendents from the maximum clade credibility tree (Figure 3A), we noticed that a small number of introductions resulted in a sustained expansion of local transmission while the remaining majority of introductions produced few downstream infections. The extent to which some individuals tend to contribute disproportionately to infection events is measured by the dispersion parameter k that quantifies transmission heterogeneity (21). Lower values of the dispersion parameter correspond to a higher degree of heterogeneity in transmission. When transmission heterogeneity is high, interventions targeting the most infectious individuals can have a considerable impact on epidemic burden. Quantifying transmission heterogeneity is hence important to guide control efforts. We thus sought to quantify mpox transmission heterogeneity using a method relying on the analysis of the size distribution of clusters of identical sequences (46).

We observed that the mean size of clusters of identical sequences decreased over the course of the epidemic (Figure 7A). We found that the timing of the decrease across locations was consistent with our estimates of *Rt* obtained from the analysis of case and sequence data (Figure 5), with larger cluster sizes observed in the US than in Europe during June 2022. Globally, the size of clusters of identical sequences ranged from 1 to 118 with 61% of sequences belonging to a cluster of size greater than 1 (Figure S7). The probability to observe a cluster of a given size is determined by the effective reproduction number *R* across the period, the degree of transmission heterogeneity measured by the dispersion parameter *k* and the fraction of infections sequenced (see Methods). Figure 7B depicts how the probability to observe a cluster of size 118 (knowing we observed 2624 clusters) is impacted by *R* and *k* assuming that 5.5% of infections were sequenced (average proportion of cases sequenced throughout the epidemic). We find that for values of the reproduction number *R* greater than 1.5, observing a cluster of identical sequences of size 118 is not unlikely regardless of the value of the dispersion parameter *k*. This is consistent with the fact that in this parameter regime, the expected mean number of offspring with identical genomes is greater than 1 so that we expect some clusters of identical sequences to not go extinct (22). For a value of the dispersion parameter similar to what has been estimated during previous mpox outbreaks (e.g. 0.36 in (23)), the reproduction number would need to be greater than 1.31 for this probability to reach 5%. Considering a lower dispersion parameter value (0.1 which is on the lower range of what has been estimated across different pathogens (21)) would still require the reproduction number to be greater than 1.21 for this probability to reach 5%. This suggests that transmission heterogeneity alone (without a reproduction number greater than 1) is unlikely to explain the size of the large polytomy observed at the beginning of the epidemic. Overall, the large first polytomy is highly consistent with a reproduction number greater than 1 at the beginning of the mpox outbreak. This aligns with reproduction number estimates obtained from our phylodynamic analysis (Figure 5), which is indicative of mpox spread within the community.

**Figure 7.**
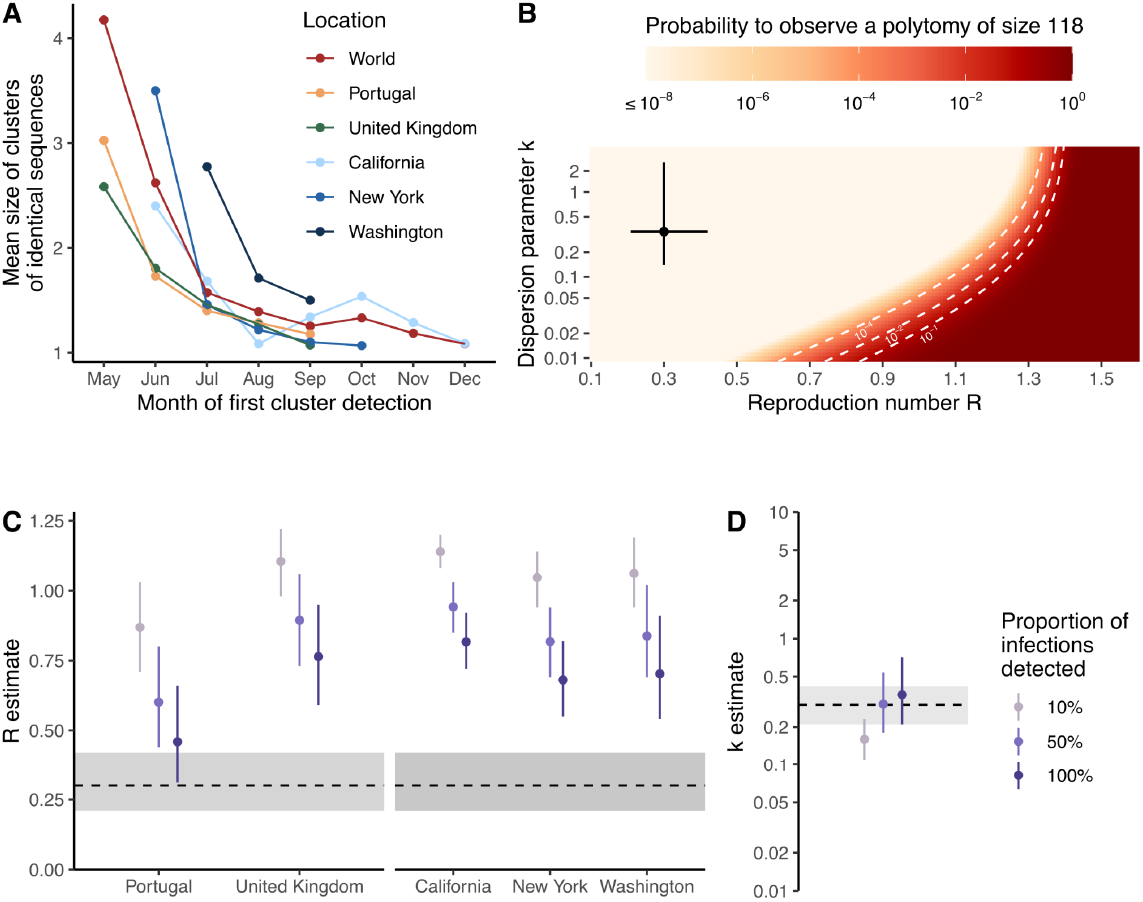
Transmission heterogeneity estimates obtained from clusters of identical mpox sequences. **(A)** Mean size of clusters of identical sequences for different geographical regions by month of first cluster detection. **(B)** Probability to observe a cluster of size 118 among 2624 clusters as a function of the reproduction number *R* and the dispersion parameter *k* assuming 5.5% of infections are sequenced. Estimates of **(C)** the reproduction number *R* by geographical unit and **(D)** the dispersion parameter *k* across geographical units from August 2022 exploring different assumptions regarding the proportion of infections detected. In B, the point corresponds to estimates obtained by Blumberg and Lloyd-Smith (43) from the analysis of epidemiological clusters during previous outbreaks. The segments correspond to the associated 95% confidence intervals. In C-D, points correspond to maximum likelihood estimates and vertical segments to 95% likelihood profile confidence intervals. The horizontal dotted line and the shaded area correspond to estimates obtained by Blumberg and Lloyd-Smith (43) from the analysis of epidemiological clusters during previous outbreaks. In B, the dotted white lines correspond to contour lines for probabilities of 10^−4^, 10^−2^ and 10^−1^.

We then estimated *R* and *k* during the decreasing phase of the epidemic in different geographical regions (Figure 7C-D, Tables S3-4). Assuming that half of infections were detected, we estimated *k* across locations at 0.30 (95% CI: 0.18-0.54) and reproduction numbers below unity across locations (Table S3). This corresponds to heterogeneity in transmission with 65% - 72% of infected individuals producing 0 offspring (and hence the remainder responsible for all transmission events). Assuming a greater fraction of infections were detected lead to lower estimates of *R* and greater estimates of *k*. This had however little impact on the fraction of individuals producing 0 offspring (Table S3). Allowing the dispersion parameter *k* to vary between locations resulted in similar estimates, though with considerably more uncertainty (Table S4). Our results suggest considerable transmission heterogeneity which could be explained by the structure of the sexual contact network in MSM (24,25). Our estimate is consistent with those previously obtained for sexually-transmitted infections spread between MSM (e.g. dispersion parameter of 0.257 estimated during a gonorrhea outbreak in MSM (26)).

## Discussion

Despite the heightened focus on public health surveillance of emerging infections since the start of the SARS-CoV-2 pandemic, MPXV sparked regional epidemics around the world, contributing to a high degree of morbidity among those affected (24,27,28). In this study, we present both a global and regional view of mpox detection, expansion, and containment by jointly analyzing genomic, mobility, and epidemiological data. We find evidence of rapid spread following initial regional viral seeding events, community transmission prior to detection by local public health surveillance, differential changes in case-detection throughout the epidemic, a limited role of viral introductions in prolonging regional epidemics, a large degree of transmission heterogeneity, and limited impact of vaccination campaigns during the early phases of the North American epidemic.

Despite double stranded DNA viruses typically exhibiting a slower evolutionary rate than RNA viruses (29), clade IIb of MPXV has been found to have a significantly faster evolutionary rate since transitioning to sustained human-to-human transmission driven by APOBEC3 editing (30). While the evolutionary rate of the variola virus (a closely related poxvirus to MPXV) has been previously estimated to be about 9× 10^−6^ substitutions per site per year (31), we infer the evolutionary rate of the B.1 lineage of MPXV to be 8. 41× 10^−5^ (95% HPD 7. 71× 10^−5^ to 9. 10× 10^−5^) substitutions per site per year or approximately 16.6 substitutions per genome per year (compared to the 1-2 substitutions per genome per year for variola virus). This increased evolutionary rate approaches the rate of many RNA viruses (32) and allows for a strong phylogenetic signal (Figures S2 and S5) to analyze epidemic spread and dynamics.

While prior studies have analyzed the global spread of MPXV via phylogenetic methods (13–15), they were often limited by small sample sizes and a superficial description of regional trends. Recent advances in phylodynamic and phylogenetic methods have been developed to tackle issues of low genetic diversity and biased sampling where phylodynamic uncertainty is reduced by the joint inference of genomic information alongside relevant predictors, such as epidemiological and mobility information (25,33–35). In the present study, we leverage these recent advances through the use of MASCOT-GLM, an approximate structured coalescent approach found to be more robust to sampling bias than traditional phylogeographic methods that allows for the integration of important predictors, notably estimated prevalence and air passenger volumes, to inform estimates of local transmission dynamics and regional viral migration.

These phylodynamic estimates, in addition to untangling global dispersion, allow us to explore changes in case detection and the impact of viral introductions on local spread on a regional level, highlighting global differences in epidemic outcomes. Despite the heightened interest in public health surveillance, we found evidence of early undetected spread in each region (Figure 3). These early undetected transmission events were often associated with the largest downstream clusters, while later viral introductions were quickly contained. Additionally, we found a strong influence of monthly predictors for the beginning months of the epidemic – May, June, July 2022 – with regards to estimating regional effective population size. The strong effect of the early monthly predictors implies the presence of significant case underreporting as the prevalence predictor in our model was not solely sufficient to inform inference of *Ne*. Despite worldwide attempts to improve public health surveillance, our study shows the limitations of current surveillance systems, promoting the need for broader routine specimen screening for a wide range of pathogens with outbreak potential.

*Rt* is a measure of transmissibility and has been widely used for monitoring changes in transmission dynamics and evaluating the impact of interventions (36). The most common methods for estimating *Rt* rely on a time series of case counts and the distribution of the generation time, and rely on the assumption of constant detection rates. (37,38). As our results suggest that case detection for mpox varied significantly in the early stages of the epidemic (Fig 3), such methods will result in biased estimates of the reproduction number and the impact of control measures for the mpox epidemic (37). In contrast to *Rt* estimates obtained solely from case counts (Figure S6A or (39)), we obtained estimates that are smaller in magnitude by relying on phylodynamic models informed by prevalence estimates and monthly predictors that account for changes in case detection. Case-based *Rt* calculations will be overestimated if case detection is increasing as the estimates capture both the true rise in infections and the rise in detection of infections. For mpox, case-detection stabilizes in August 2022 (Fig. 3D), after which time we expect case-based Rt estimates to be more accurate. This suggests that our approach of integrating multiple data sources would provide a more accurate estimation of mpox transmissibility and of the impact of interventions, especially in the beginning stages of an epidemic where accurate knowledge of *Rt* can have high impact in informing public health action.

An outstanding question raised during the beginning of the mpox epidemic that remains unclear is the potential impact of interventions in preventing and controlling spread (40). Similar to the early phases of the SARS-CoV-2 pandemic, the MPXV epidemic prompted considerations around travel bans and restrictions in an attempt to curb transmission to previously unaffected areas. While travel bans were ultimately not implemented, the CDC issued a series of travel recommendations and warnings for both individuals exposed to MPXV and for those traveling to areas with a high number of mpox cases on June 6, 2022 (41). Despite these travel recommendations, our models show that there were already many introduced lineages circulating in North America before June 6th (Figure 3), limiting the impact and effectiveness of these recommendations on curbing disease spread. Our results show that following initial viral seeding, viral introductions played a limited role in promoting local transmission, accounting for less than 15% of new cases in any given region studied (Figure 4). We also found that removing the influence of introductions also would have had limited impact in the timing of North American *Rt* dropping below one (Figure S6). Together this suggests little potential impact of travel restrictions after mid-May 2022 once MPXV had already been established in the region. Our estimates of transmission heterogeneity, where we found that only 28-35% of infected individuals were responsible for all transmission events observed during the decreasing phase of the epidemic (Figure 7), promotes tailoring public health interventions to high risk groups rather than population-wide policies.

We also examined the impact of vaccination campaigns on controlling the mpox epidemic in North America by comparing changes in local transmission as measured by *Rt* to the cumulative percentage of high-risk individuals in the US with vaccine-derived immunity (Figure 6). While even a half vaccination dose has been found to be effective at providing robust immunity against mpox (42,43), there was concern over the delayed start of vaccination campaigns in the US. We find that local transmission in North America decreased below one in mid-August 2022 before 10% of the population had any vaccine-induced immunity. In the present analysis, we only accounted for vaccine-derived immunity. However, we can attempt to account for immunity derived from natural infection by comparing mpox cases to the size of the at-risk population. Doing so, we find that less than 2% of the high risk MSM population in the US had reported cases of mpox as of Nov 22, 2023 (see *Methods* under *Data Sources)*. Converting this crude cumulative incidence into an estimate of the total proportion of the population infected requires knowing the reporting rate of mpox infections. If we assume complete reporting then we expect just 2% cumulative incidence, which should have negligible impact on lowering epidemic Rt. However, if the reporting rate was 10% then we expect approximately 20% cumulative incidence, which starts to have an impact on Rt. While we were unable to find a precise estimate of reporting rate in the US, prior studies in Portugal (44) and in North Carolina (45) estimate the rate of detection to be 62% (95% CI, 43%-83%) and 66% (95% CI, 44%-91%). If we assume that the US reporting rate falls on the lower bound of those estimates, we expect 4.7% cumulative incidence. Thus, in general we expect that natural immunity will have played a minor role in reducing epidemic Rt compared to behavioral modification and vaccine-derived immunity.

The incongruent population definitions (high-risk population in the US for vaccine-derived immunity and a single *Rt* estimate for the US and Canada combined) could conceivably bias our conclusions regarding the relationship between Rt and vaccine coverage. Despite the lack of publicly available vaccination information for the whole of Canada, vaccination data for Montreal (46) and Ontario (47) show that pre-exposure vaccination began at a similar time, if not later, than the vaccination efforts in the US (immunization in Montreal began on May 27, 2022 and in Ontario on June 9, 2022 compared to May 22, 2022 in the US (4)). We believe that the similar, if not later, start of vaccination campaigns in Canada biases our results in a conservative and limited fashion compared to what would be expected if the Canadian vaccination efforts began earlier than those in the US. In addition, the Canadian MSM population is estimated to be 10% of the US population (48,49), further suggesting that vaccination in Canada should have a limited role in reducing mpox *Rt* in North America. More broadly, our estimates of *Rt* and vaccine-derived immunity aggregate across large spatial regions. Further spatially resolved analyses could provide additional information about the relationship between *Rt* and vaccine coverage.

Mpox in the US and Canada spread predominantly among high-risk MSM populations (50), suggesting that the majority of the North American sequences in our study were derived from a similar (but not identical) population as used to estimate vaccine coverage. Our conclusions are concordant with those from the CDC which also found that *Rt* fell below one in August 2022 when only about 1.3% of the high risk population in the US had any vaccine-induced immunity (51). Similarly, modeling of mpox in Washington D.C. suggests that behavioral modifications within the MSM community were the main contributing factor to slowing initial mpox spread, but that vaccination campaigns were ultimately needed to definitively curb the local epidemic and prevent future outbreaks (52,53). A UK-based modeling study focusing on men who have sex with men found that vaccination could not explain the drop in mpox incidence in the region, but rather attribute the declining incidence to changes in behavior within the same community (54). Together, these findings highlight the significant effect of behavioral change among men who have sex with men in curbing the epidemic as well as emphasize the need for prompt public health response in order to maximize the population-level effectiveness of vaccination campaigns.

In conclusion, our study integrates diverse data sources to provide novel insights on the spread and control of mpox. Despite global efforts to improve molecular surveillance, our study shows that early unrecognized spread was critical to driving the initial epidemic. Once the mpox epidemic was recognized, behavioral modification in the MSM community resulted in a sharp decline in *Rt* in North America ahead of vaccination rollout in the US. Our findings are relevant for policymakers in promoting broader routine specimen screening as a core tenant of pandemic preparedness. Recent emerging disease outbreaks – Zika, Ebola, SARS-CoV-2, and now mpox – have been characterized by late public health detection and cryptic local transmission as a result (55–57). Our work shows that rapid pathogen detection and subsequent behavioral change could be sufficient to curb epidemic spread. Additionally, our work prompts swift public health investments and interventions to protect marginalized and vulnerable populations from mpox and other emerging infections (11,28).

### Limitations of the study

Our study has noteworthy limitations. Our genomic data from GenBank only cover a small selection of countries and regions, suggesting that we are missing transmission events that involve unsampled countries especially from regions such as Asia, Oceania, and Africa, although mpox cases in these areas in summer 2022 were limited and unlikely to significantly impact our results. Additionally, the changing availability of genomic sequencing, as well as unequal sampling across the regions study affect the probability that a case shows up as a sequence in our dataset through the period studied. If viruses migrate frequently between our study countries and countries that lack genomic sampling, the lack of samples that might interdigitate with samples from the study country may affect our ability to distinguish separate introductions. Despite this potential bias, the 2022 mpox epidemic mainly affected Europe and the Americas, which are regions that are well represented in our study, limiting the effect of this bias. Additionally, we attempted to account for this variation by weighting the subsampling for our phylogeographic (DTA) analysis according to confirmed case counts, and by oversampling undersampled regions (and downsampling overrepresented regions) in our MASCOT-GLM analysis (Figure S1) as well as by adding in estimated prevalence as an predictor in the model in an effort to account for this variation.

Bayesian coalescent models assume random sampling of infected individuals, meaning that targeted sampling of superspreader events, or via contact tracing, could bias our phylodynamic estimations. We attempt to quantify the extent of transmission heterogeneity via our estimates of overdispersion (Figure 7). In the analysis of transmission heterogeneity, we explicitly accounted for the fraction of cases sequenced and explored several assumptions regarding the proportion of infections detected by the surveillance system. This was done assuming that all infections had the same probability of being detected as cases and sequenced. Active surveillance targeting larger clusters could lead to underestimating the extent of transmission heterogeneity (23,58).

We see a discrepancy in the TMRCA between various models (Table S2) and find our estimates to be highly dependent on the tree prior and thus should be interpreted with caution. Inference of TMRCA is dependent on the estimate of effective population size in early 2022. Different tree priors assume different parametric forms of effective population size and so differ in TMRCA estimates. The rapid exponential growth observed in early 2022 suggests that effective population size should be low in January-March 2022. This information is used by the DTA skyline and skygrid models, as well as the MASCOT-skyline model, resulting in TMRCA estimates close to the earliest March sequences. Consistently, the MASCOT-GLM model estimates the coefficient of the monthly predictor for April 2022 and earlier at -1.09 (95% HPD: -1.89 – 0.00, Fig. 3D), again supporting a small effective population size in this time period. We suggest a conservative interpretation of these results supporting a TMRCA between September 2021 and March 2022.

## STAR Methods

### Resource availability

#### Lead contact

Further information and requests for data should be directed to and will be fulfilled by the lead contact, Miguel I. Paredes.

#### Materials availability

This study did not generate new unique reagents, but raw data and code generated as part of this research can be found in the Supplemental Files, as well as on GitHub as specified in the Data and Code Availability section below.

#### Data and code availability

This paper analyzes existing, publicly available data. Curated sequence data, Nextstrain builds, BEAST XMLs, scripts, sequence information, and de-identified data can be found at https://github.com/blab/mpox-dynamics. We provide an acknowledgements table for all sequences used in the manuscript in this repository.

## Method Details

### Genomic data and maximum likelihood tree generation

All available MPXV sequences were downloaded from GenBank while excluding sequences from countries with five or fewer sequences, leaving Austria, Belgium, Canada, Colombia, France, Germany, Italy, Peru, Portugal, Slovakia, Slovenia, Spain, Switzerland, United Kingdom, and the USA. Sequences with ambiguous date of collection in the month column, with a sample collection earlier than January 2022, and flagged as being low quality by Nextclade https://docs.nextstrain.org/projects/nextclade/en/stable/user/algorithm/07-quality-control.html)(60) were excluded. Given that the 2022 epidemic was found to be driven by MPXV clade II, lineage B.1 (14,61), any sequences not part of lineage B.1 were also excluded, resulting in 3013 genome sequences included in our analysis.

A temporally-resolved phylogeny was created using a modified version of the Nextstrain (62) monkeypox workflow (https://github.com/nextstrain/monkeypox), which aligns sequences against the MPXV_USA_2021_MD (accession ON918611) reference using nextalign (60), infers a maximum-likelihood phylogeny using IQ-TREE (63) with a GTR nucleotide substitution model, and estimates molecular clock branch lengths using TreeTime (64). The resulting phylogeny specific to this dataset can be found at https://nextstrain.org/groups/blab/monkeypox/hmpxv1.

### Regional geographic scales

Due to the low number of sequences from various countries, we analyzed mpox spread at the scale of global regions. We focused on five regions with the highest number of publicly available sequences on Genbank: Central Europe, North America, South America, Southern Europe, and Western Europe. Country to region mapping can be found in Table S1.

### Data Sources

Data on the number of reported mpox cases per region per month were downloaded from OWID (https://ourworldindata.org/; last accessed on February 13 2023).

Population sizes for each country were downloaded from the World Bank (https://data.worldbank.org/indicator/SP.POP.TOTL) and aggregated based on respective countries and then regions as described in the previous section.

To compare vaccination rates with changes in *Rt*, we accessed publicly available vaccination counts from the CDC (https://www.cdc.gov/poxvirus/mpox/response/2022/vaccines_data.html) as well as the cumulative percentage of high risk individuals vaccinated (51). The CDC defined “high-risk individuals” as MSM for whom preexposure prophylaxis against infection for HIV is clinically indicated as well as MSM who are living with HIV. In order to account for the development of immunity, we followed the CDC method of assuming the development of immunity took two weeks following vaccination (51) and thus only considered individuals as “having vaccine-induced immunity” after reaching two weeks from the date of first vaccination. We additionally estimated that 1.88% of the high risk population in the US was infected with mpox (calculated by dividing the total number of confirmed mpox infections in the US, which is 31,010 as of November 22, 2023, by 1,647,121, which is the total number of US individuals estimated by the CDC to be at high risk for mpox infection.)

We used air travel data from the International Air Transport Association (IATA) quantifying the monthly number of passengers on origin-destination itineraries between airports in the 15 included countries (65).

#### Site masking

We found that fewer than 1% of nucleotide positions out of 197,209 total sites in the MPXV sequence alignment were phylogenetically informative, ie. polymorphic. To reduce computational runtime for phylogeographic reconstruction (discrete trait analysis), we masked 90% of invariant positions from the MPXV alignment prior to further analysis. The Nextstrain monkeypox workflow produces a BED file containing phylogenetically uninformative or misleading alignment positions to be masked. A VCF file was generated from the alignment using SNP-sites v2.5.1 (66). We identified variable positions from the VCF using Pysam v0.20.0 (67). Next, we selected a random subset of 90% of all invariant positions to remove and appended the remaining nucleotides to the BED file. A new alignment of 19,721 positions was generated with the modified BED file using the Nextstrain workflow. Clock rate estimates inferred with a masked alignment were adjusted by a magnitude of 10 in order to account for the degree of masking.

#### Phylogeographic analysis

To investigate the dispersal history of MPXV among five global regions, we first conducted an asymmetric discrete trait phylogeographic analysis (68) using the Bayesian stochastic search variable selection (BSSVS) model implemented in BEAST 1.10 (69). For this analysis, we considered each global region as a discrete location and employed subsampling weighted by mpox case counts for each region using a random seed, resulting in a final subset of 1004 sequences (distribution across countries and regions shown in Supplementary Table 1). We masked the alignment as described above. We employed a strict molecular clock with a uniform distribution from 0 to 1 and an initial value of 6× 10^−5^ and a GTR+Γ nucleotide substitution model. We used a Skygrid coalescent tree prior allowing grid points to change every two weeks (70). Two independent Markov chain Monte Carlo (MCMC) procedures were run for 5× 10^8^ iterations and sampled every 1000 iterations. Resulting posterior distributions were combined after discarding initial 20% of sampled trees as burn-in from each of them. We used Tracer 1.7 (71) to assess convergence and to estimate effective sampling size (ESS). These values were all >150. We then used TreeAnnotator 1.10 to obtain a maximum clade credibility (MCC) tree removing the first 20% of iterations for burn-in.

The number of viral imports and exports between regions was estimated by calculating the number of regional transitions walking from tips to root in the posterior set of trees and calculating the median as well as the 50% and 95% highest posterior density estimates (HPD). Following Bedford et al. (72), persistence time was measured by calculating the average number of days for a lineage to leave its sampled region, walking backwards up the phylogeny from the tip up until the node location was different from the tip region.

In a secondary analysis, in order to check the accuracy of ancestral state reconstructions as well as the strength of genomic signal, out of the same 1004 sequences, 10% had their locations masked and then reconstructed (55) via the same discrete trait analysis described above.Reconstruction accuracy was assessed by comparing the most likely reconstructed location with the true location.

### Estimation of mpox incidence, prevalence, and effective reproduction number via case counts

To jointly estimate mpox case incidence, prevalence, and effective reproduction number, we used the renewal equation framework from Figgins and Bedford (73). The time-varying effective reproduction number (i.e. the average number of secondary cases infected by a single primary case) was modeled using a 4th order spline with 5 evenly spaced knots assuming a discretized gamma-distributed generation time with mean 12.6 days and standard deviation 5.7 days (6). Case counts were modeled using a zero-inflated negative binomial distribution. This model produces posterior estimates of daily incidence (defined as the number of newly infected individuals in absolute counts) and effective reproduction number. We then used this incidence and an assumed gamma-distributed infectious period with a mean of 4.5 days to compute the prevalence, which we define as the number of actively infected individuals in absolute counts (6).

Models were fit to aggregated case counts for each region using full-rank stochastic variational inference. Optimization was performed using the ADAM optimizer with learning rate 4e-3 and for 50,000 iterations and 500 samples were drawn from the approximate posterior.

### Estimated importation intensity

We estimated the monthly importation intensity of mpox between the five selected global regions between May and December 2022 using air travel data, estimated regional prevalence and regional human population size. The monthly estimated importation intensity (EII) is an estimate of the number of mpox cases imported into each region during a given month, calculated as

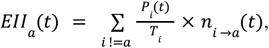

where EII for region *a* at month *t* is computed using the estimated mpox prevalence *P*_*i*_(*t*) in a different region *i*, the population size *T*_*i*_ in region *i* and the number *n*_*i* →*a*_(*t*) of air passengers traveling from region *i* to region *a* (adapted from Fauver et al.(74)). The sum over every global region excluding domestic travel. We used the prevalence estimates obtained from case data as described in the previous paragraph.

### MASCOT-GLM

To analyze the transmission dynamics within and between each global region, we used an adapted version of MASCOT (18). MASCOT is an approximate structured coalescent approach (75) that models how lineages coalesce (share a common ancestor) within the same locations and migrate between locations. We used generalized log-linear models (33) to estimate whether estimated regional mpox prevalence and air passenger volumes are predictive of MPXV effective population sizes and migration rates over time, respectively. Additionally, in order to account for differential underreporting by month, ten additional effective population size predictors were added, one for every month of the time period studied from April 2022 through January 2023. Empirical predictors were obtained via data sources described above. The model included error terms to account for observation noise and omitted predictor variables. We implemented a MASCOT-GLM (33) analysis with BEAST2 (76) software, allowing the effective population sizes and the migration rates to change every week. We performed effective population size and migration rate inference using an adaptive multivariate Gaussian operator (77) and ran the analyses using an adaptive Metropolis-coupled MCMC(78) using four chains with a length of 2. 5× 10^8^. For this analysis, we employed equal temporal subsampling to enrich for undersampled regions by randomly choosing a max of 11 sequences per region per calendar month via Augur filter (79), resulting in 587 included sequences. We chose an equal temporal subsampling scheme due to recent work showing that maximizing spatiotemporal diversity reduces bias in MASCOT-GLM (19) The unmasked alignment was used for all MASCOT-GLM analyses.

### MASCOT-Skyline

In order to investigate the degree of genomic signal and influence of empirical predictors on tree reconstruction, we reran our MASCOT analysis without empirical predictors using a MASCOT-Skyline approach. To allow for population sizes to change over time, we modeled the effective population sizes similar to the Skygrid approach for unstructured populations (70). We estimated the effective population size for each location between time *t*=0×tree height, …, *t*=1×tree height. Between each time point where we estimated the *Ne*, we assumed exponential growth. We assume the prior on the effective population size over time to be a Gaussian Markov random field (GMRF) and estimate the variance of the GMRF prior on the effective population size over time. We assume the GMRF prior for each state to have the same varianc We assumed the migration rate to be constant forward-in-time, 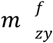, between states *y* and *z*. As the structured coalescent assumes backwards-in-time migration rates, we assumed that the backwards-in-time rate of migration between state *y* and z, 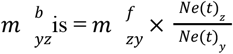. To infer effective population sizes and migration rates over time, we employed an adaptable multivariate gaussian operator (77).

#### Posterior processing

Parameter traces were visually evaluated for convergence using Tracer, tree distributions were visually inspected using IcyTree (80), and 20% burn-in was applied for all phylodynamic analyses. All tree plotting was performed with baltic (https://github.com/evogytis/baltic) and data plotting was done using Altair (81).

The number of migration events between regions was estimated by calculating the number of regional transitions walking from tips to root in the posterior set of trees and calculating the median as well as the 50% and 95% highest posterior density estimates (HPD). In order to calculate the migration rate for each model, we divided the total migration count for each tree in the posterior set by the tree length (the sum of all branch lengths) and then calculated the mean and 95% HPD.

#### Estimating percentage of new cases due to introductions

We estimated the percentage of new cases due to introductions for each global region by adapting the methods previously described (34,82). The percentage of cases due to introductions π at time *t* can be calculated by dividing the number of introductions at time *t* by the total number of new cases at time *t*. We first represented the total number of new cases in a region as the sum of the number of introductions and the number of new local infections due to local transmission, resulting in the following equation:

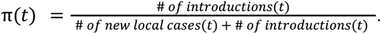

We estimated the number of new local cases at time *t* by assuming the local epidemic in each global region follows a simple transmission model, in which we derived the number of new cases at time *t* as the product of the transmission rate β (new infections per day per individual) multiplied by the number of people already infected in that region *I*. For the number of introductions, we similarly assumed that the number of introductions equals the product of the rate of introduction (introductions per day per infectious individual, which we refer to as migration rate *m*) and the number of people already infected in that region *I*. We use the number of infected individuals in the destination region rather than the origin region for calculating the number of introductions since the approximate structured coalescent approach models epidemic processes as backwards-in-time, resulting in the equation containing only information about the number of infected individuals in the destination region (more information on backwards migration rates below). We then rewrote the above equation as

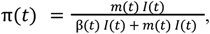

where *I*(*t*) denotes the number of infected people in that region at time *t*. Given the presence of\ *I*(*t*) in every element, we factored out *I*(*t*) to arrive at

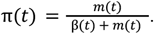

For each region, we considered introductions at time *t* to be the sum of the introductions coming into the region from each other global region, assuming a negligible number of introductions from unincluded regions. We define the percentage of new cases due to introductions π at time *t* for region *y* as

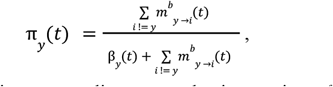

where *m* _*i* →*y*_ denotes the migration rate per lineage per day into region *y* from every other region.

In a SEIR transmission modeling framework (employed due to the incubation period of MPXV), the transmission rate β is a function of the infectious period γ, the incubation period σ, and the exponential growth rate *r (*as adapted from Example 4 in Ma 2020 (83)):

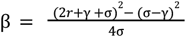

To compute the growth rate in region *y*, we assumed that differences in effective population size between adjacent time intervals can approximate the growth rate *r* and thus 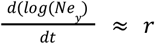. In addition, we assumed that 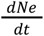 is independent from the rate of introduction. We calculated the growth rate of the effective population size 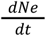 as

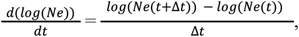

where *Ne*(*t*) denotes the effective population size of a region at time *t*. We ran our MASCOT-GLM analysis using weekly time intervals but averaged over three week intervals (Δ*t* = 3) for the growth rate in order to reduce noise and account for the long generation time for mpox.

By also assuming an expected time until becoming uninfectious γ for each individual of 4.5 days and an incubation period σ of 8 days (6), we calculated the transmission rate β at time *t* in region *y* as

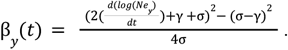

Since the coalescent, which MASCOT approximates, works backward-in-time, we calculated the rate of introductions into each global region *m*_*y*_(*t*) as the backwards migration rate *m* ^*b*^ _*y*_ (*t*) from inferred MASCOT parameters. To compute the backwards migration rate, we extract the forward-in-time migration rate *m*^*f*^ _*yi*_ (*t*), where *i* refers to a different region in a combination of global regions *c*, that is inferred via MASCOT-GLM, and then calculate the backwards-in-time migration rate into region *y*, as the sum of the products of the ratio of effective population sizes 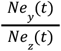 and the forward migration rates:

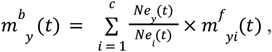

where *Ne*_*y*_ (*t*) refers to the effective population size in region y at time *t* and *Ne*_*i*_ (*t*) refers to the effective population size in a different region *i* from a combination of global regions *c* at time *t*.

### Estimating the effective reproductive number Rt from pathogen genomes

We calculated the effective reproductive number *Rt*, the time-varying average of secondary infections from a primary infected individuals, in each region, assuming an exponentially distributed infectious and incubation period of mean respectively 1/γ and 1/σ, yielding 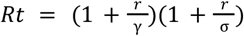 (84). Additionally, we sought to separate out the contributions of introductions versus local transmission to *Rt*_*t*_ in each region. To do so, we modified the *Rt* equation to include the percent of new cases from introductions as an estimate of local community spread so that 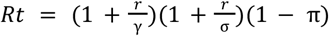, where π refers to the percentage of new cases due to introductions as described above.

### Estimating transmission heterogeneity

We analyzed the size distribution of clusters of identical mpox sequences to characterize the disease’s offspring distribution (22). We assumed that the offspring distribution follows a negative binomial distribution characterized by its reproduction number *R* and its dispersion parameter *k* (21). The probability *r*_*j*_ that a cluster of identical sequences of size *j* can be derived as

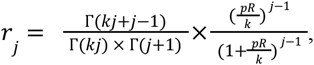

where *p* denotes the probability that a transmission event occurs before a mutation event.

In practice, only a fraction of infections are sequenced. The probability 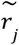 to observe a cluster of size *j* was thus derived as:

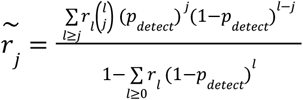

where *p*_*detect*_ denotes the fraction of infections sequenced.

The probability for an observed cluster of identical sequences to be of size at least *J* can then be computed as 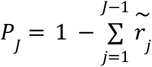. The probability to observe at least a cluster of size *J* among *n*_*clust*_ clusters is thus equal to 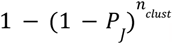.

Former work has shown that the size distribution of clusters of identical sequences can be used to infer the reproduction number and the dispersion parameter when the mean number of offspring with identical sequences lies below 1 (22). For the 2022 mpox epidemic, this would correspond to values of the reproduction number lying below 1.5 (22). To ensure this criterion was met, we analyzed the size distribution of clusters of identical mpox sequences for different geographical units (Portugal, the United Kingdom and US states California, New York and Washington) from August 2022, which corresponds to the decreasing phase of the epidemic (Figure S7). We generated the size distribution of clusters of identical mpox sequences for these different geographical units and defined clusters temporally based on their date of first detection. We estimated the fraction of cases sequenced in these different regions from August 2022 by computing the ratio between the number of sequences used and the number of cases publicly reported. We first inferred the parameters of the offspring distribution assuming that the dispersion parameter was the same across these geographical units and estimating a reproduction number for each of these geographical units. This was done by considering different assumptions regarding the fraction of infections detected (10%, 50% and 100%) and assuming a probability that transmission occurs before mutation equal to 66% (84). We also ran a location-specific model and estimated the reproduction number and the dispersion parameter for these each region. We assumed that clusters of identical sequences stemmed from local transmission dynamics. This hypothesis is supported by the small contribution played by introductions estimated from the phylogeographic analysis.

We also generated the distribution of cluster sizes worldwide. We explored how different assumptions regarding *R* and *k* impacted the probability to observe a cluster of size 118 (the largest cluster observed) among the 2624 clusters of identical sequences observed. This was done assuming that 5.5% of infections were sequenced (which corresponds to the fraction of cases sequenced since the beginning of the epidemic).

## Supporting information

Acknowledgement tables for sequences used in study

## Acknowledgments

We would like to thank Allison Black for constructive feedback, discussions, and edits. We gratefully acknowledge all data contributors, i.e. the Authors and their Originating laboratories responsible for obtaining the specimens, and their Submitting laboratories for generating the genetic sequence and metadata and sharing via GenBank. The laboratories and institutions that contributed more than ten sequences for this study are as follows: Los Angeles County Public Health Laboratories, Los Angeles County Department of Public Health, UW Virology, Laboratory Medicine, UKHSA, Research and Evaluation, CDC, DHCPP-PRB, Robert Koch Institute, Centre for Biological Threats, Highly Pathogenic Viruses, National Institute of Health Doutor Ricardo Jorge, Portugal (INSA), Department of Infectious Diseases, Laboratorio Departamental de Salud Publica de Antioquia, Antioquia, Instituto Nacional de Salud, Direccion de Investigacion en Salud Publica, National Microbiology Laboratory, Public Health Agency of Canada, Institute National de Saude Doutor Ricardo Jorge (INSA), Portugal, CDPH, VRDL, IHU - Mediterranee Infection, MEPHI, Centre for Biological Threats, Highly Pathogenic Viruses, Robert Koch Institute, Germany, New Jersey Department of Health, Public Health and Environmental Laboratories, Rush University Meical Center, Regional Innovative Public Health Laboratory (RIPHL), University of Nebraska Medical Center, Environmental, Agricultural, and Occupational Health, Universidad Tecnologica de Pereira, Laboratorio de Biologia Molecular y Biotecnologia / Facultad de ciencias de la salud, Royal Infirmary of Edinburgh, Viral Genotyping Reference Laboratory, Institute of Microbiology and Immunology, Faculty of Medicine, University of Ljubljana, Laboratory for Diagnostics of Zoonoses and WHO Centre, Institute of Tropica Medicine, Department of Clinical Sciences, Medical University of Vienna, Center for Virology, Instituto Nacional de Salud Peru, Laboratorio de Biotecnologia y Biologia Molecular. We have included a detailed acknowledgements table in Supplementary Data.

## Author Contributions

Conceived and designed the study: MIP, NA, TB

Curated the data: MIP, NA, VC, CTK

Conducted the analysis: MIP, NA, MF, VC, CTK, NFM

Advised on analysis: VC, PL, JTM, TB

Drafted the manuscript: MIP, NA, MF, NFM, CTK

Reviewed and edited the manuscript: All authors

### Ethics Approval

All data used in this study is publicly available, suitably anonymized viral sequence data. As such it does not constitute human-subjects research.

## Data Availability

Nextstrain builds, BEAST XMLS, scripts, sequence information, and de-identified data can be found at https://github.com/blab/mpox-dynamics. All sequences are available on GenBank with accession numbers found in the supplementary information.

## Supplementary Information

**Figure S1:**
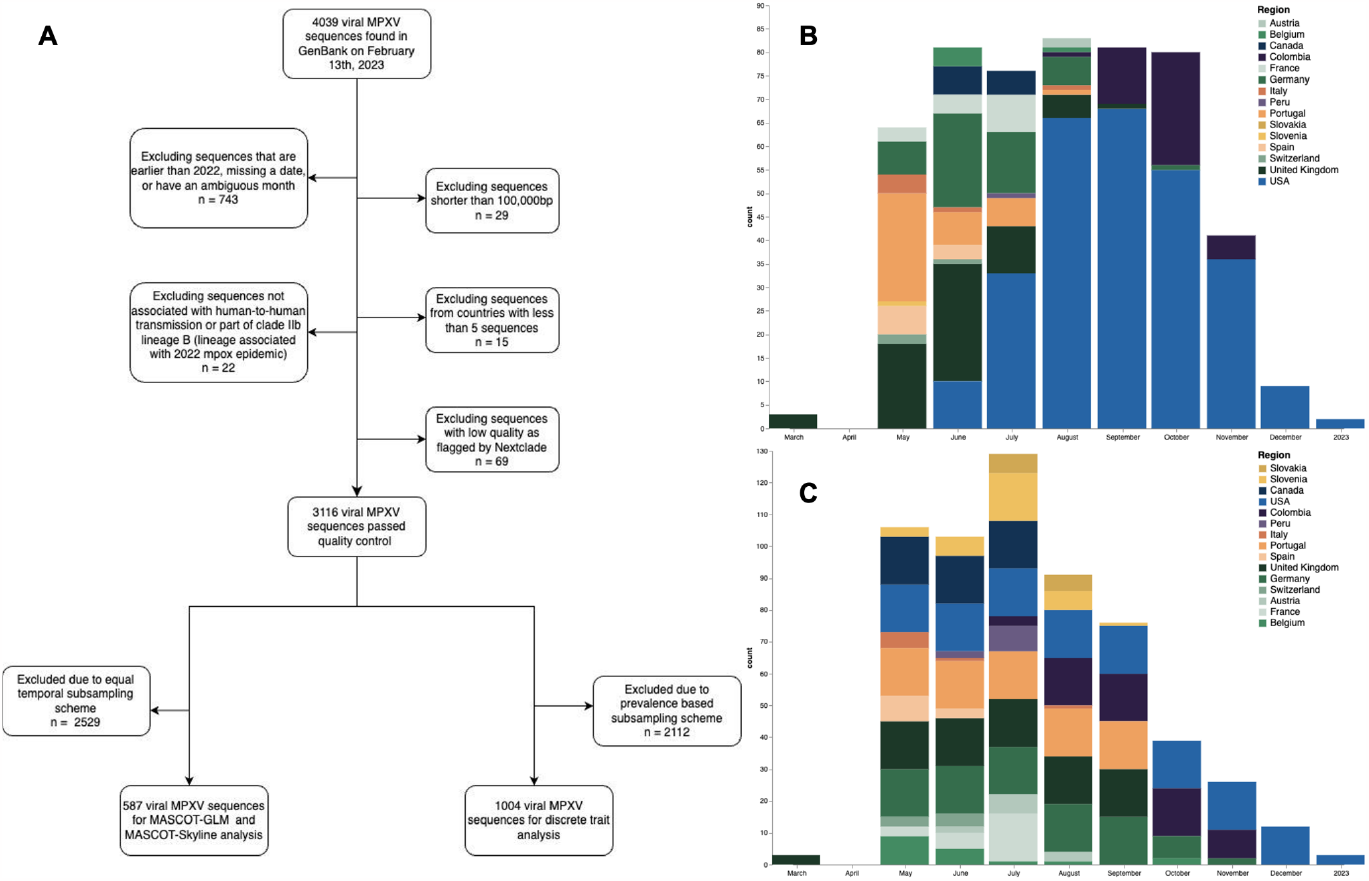
Subsampling for phylogeographic and phylodynamic inference, related to Figure 1. **(A)** Flow diagram displaying the inclusion and exclusion criteria for the final two analytic samples **(B)** Temporal Distribution of 1004 genomes used for phylogeographic analysis. Genomes were subsampled using confirmed case counts as weights. **(C)** Temporal distribution of 587 genomes used for MASCOT-GLM analysis. Subsampling was done to promote an equal number of samples from each deme for each month in order to oversample underrepresented countries.

**Figure S2:**
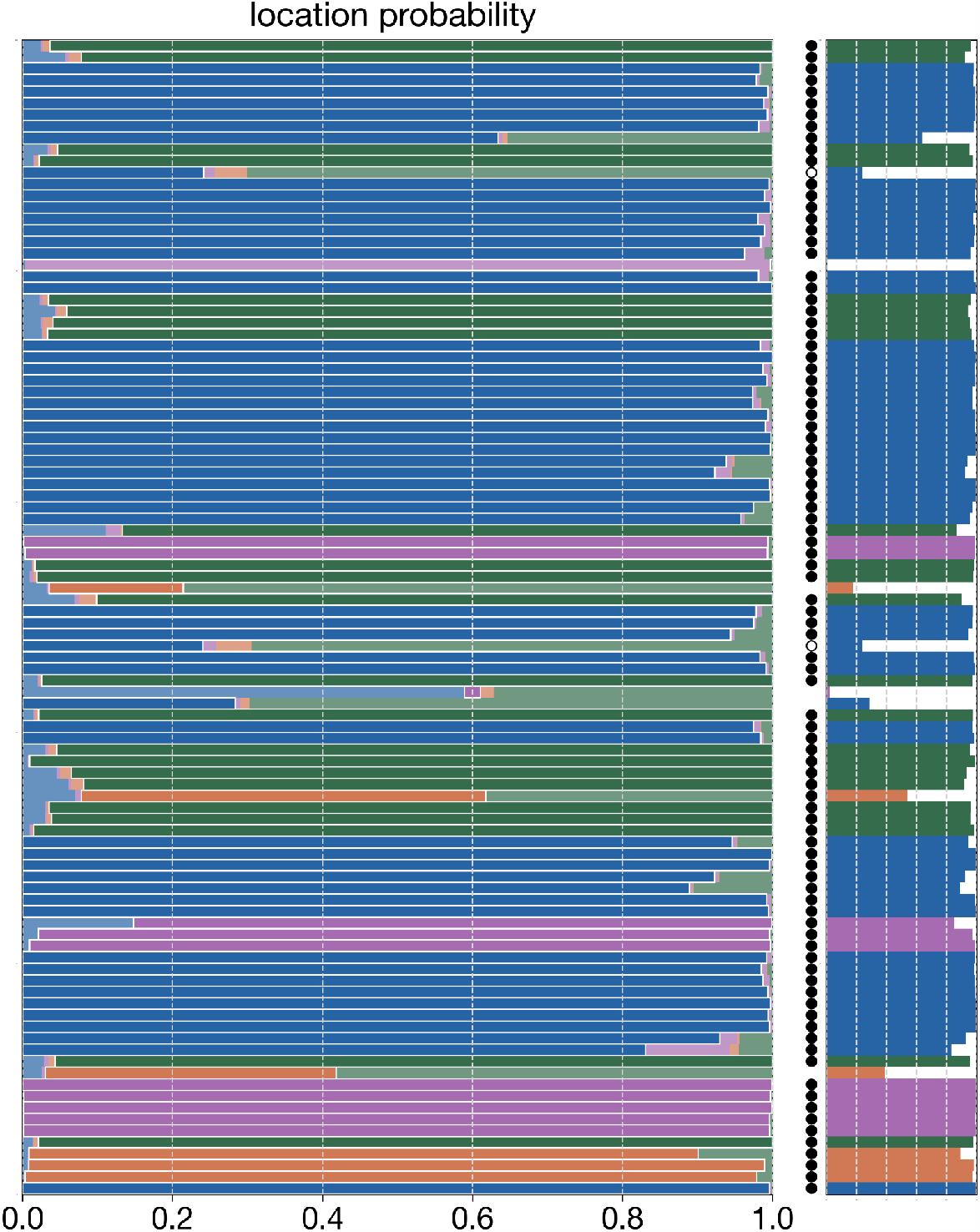
Masked tip location inference, related to Figure 2. Horizontal bars indicate the posterior distribution of masked tip locations, coloured by region.. The correct location of each tip is outlined in white with the smaller plot to the right showing only the posterior probability of the correct location. Bars marked with an open circle indicate cases where the correct location is within the 95% credible set and solid circles indicate cases where the location with the most probability mass is also the correct location.

**Figure S3.**
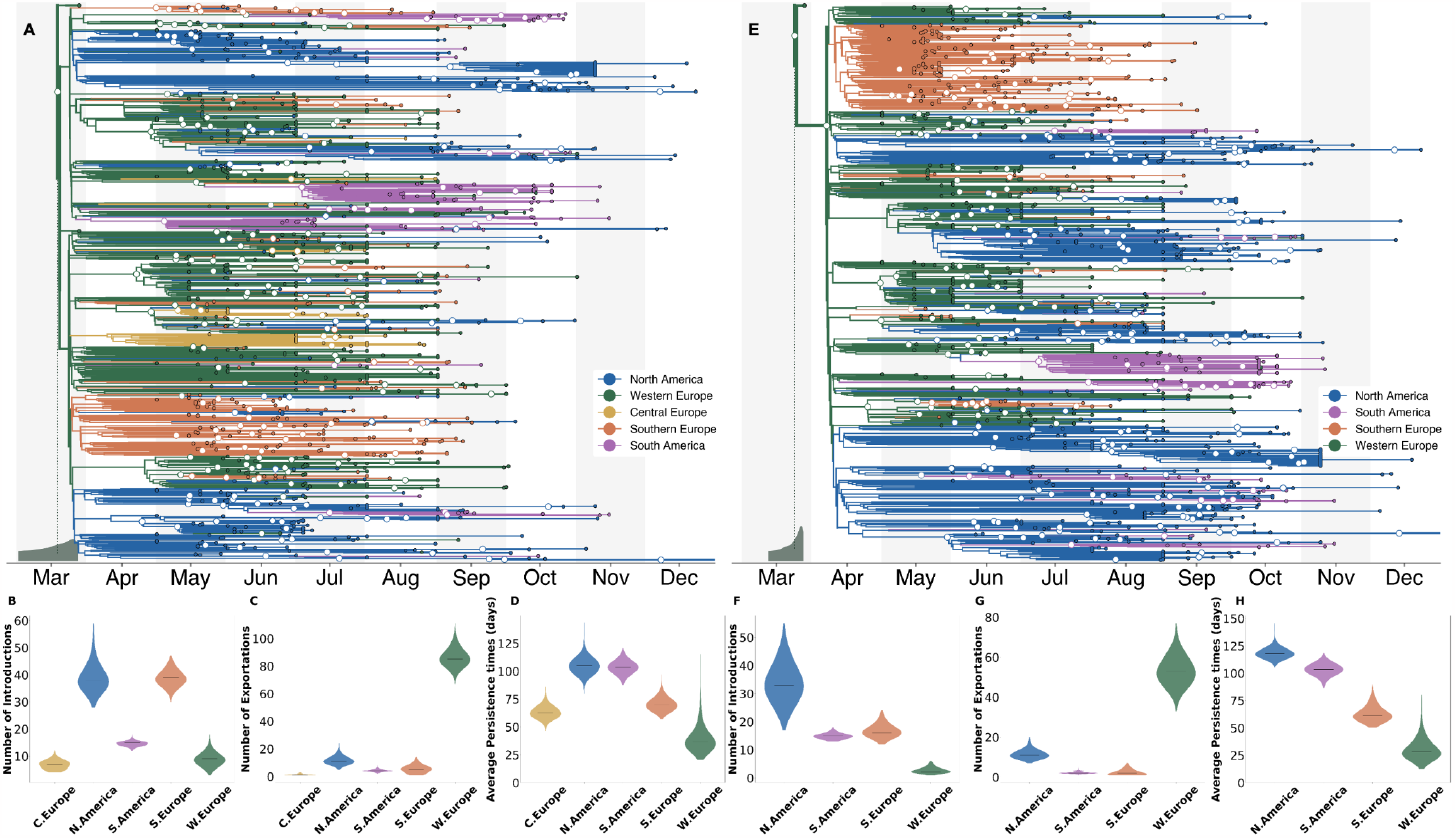
Phylogeographic analysis using alternative subsampling schemes, related to Figure 2. In comparison with main Figure 2 which uses a case-count-based subsampling scheme, **A-D** used an equal spatiotemporal subsampling scheme which attempted to sample an equal number of sequences from each region for each year-month, which is the same strategy used for the MASCOT-GLM analyses. **E-H** repeat the analyses but sampled sequences directly from global regions irrespective of country of origin. (**A & E**) The maximum clade credibility tree summary of the Bayesian inference conducted using asymmetric discrete trait analysis and Skygrid prior on 991 (**A**) and 1019 (**E**) sequences. Colors correspond to the regions in the legend. Ancestral nodes with greater than 50% posterior support are highlighted with a white circle overlaid. Inset histogram on bottom left corner shows 95% interval for the time to most recent common ancestor (TMRCA)(**B-D & F-H**) Estimated number of introductions (**B & F**), exports (**C & G**), and average time of local persistence in days (**D & H**) for each global region. Horizontal black line denotes median estimates.

**Figure S4:**
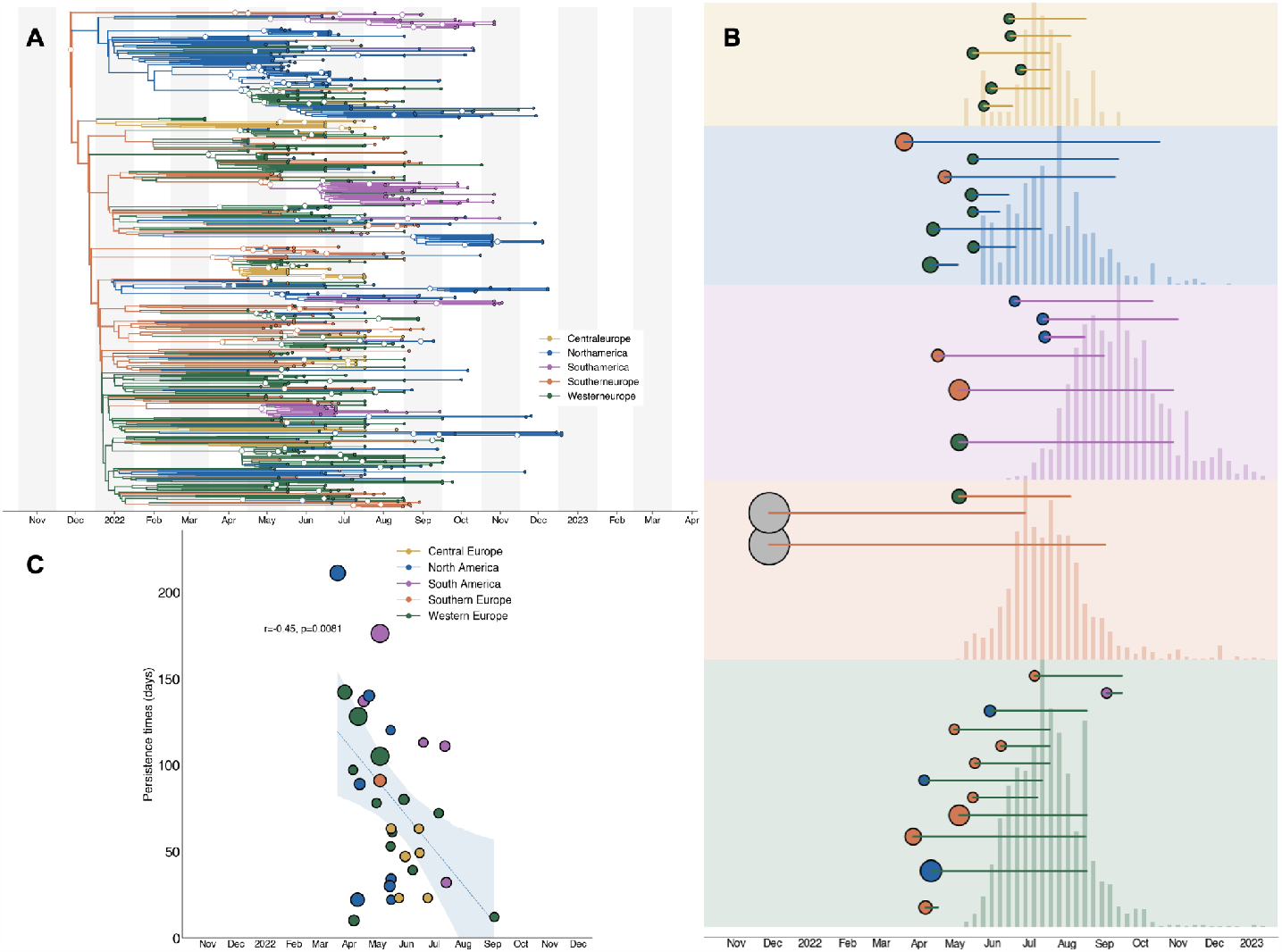
Analysis of introductions inferred via MASCOT-GLM, related to Figure 3 and 4. **(A)** The maximum clade credibility tree summary of the Bayesian inference conducted using MASCOT-GLM on 587 sequences. Colors correspond to the regions in the legend. Ancestral nodes with greater than 50% posterior support are highlighted with a white circle overlaid. **(B)** Exploded subtrees for each region with only the introductions with greater than 50% posterior support showing that early underdetected introductions lead to longer transmission chains. Color at introduction origin represents inferred source region and size of the circle at the origin is proportional to the number of downstream tips. Length of line coming out of each introduction origin represents the length of the transmission chain. Case counts are overlaid for each region. **(C)** Relationship between estimated date of introduction and persistence time with the first two large introductions removed. Each circle represents a single viral introduction with greater than 50% posterior support into the region denoted by the color (i.e. a green point represents an introduction into Western Europe). The size of each point is proportional to the size of the outbreak cluster resulting from each introduction with larger circles representing more resulting downstream tips. Blue dashed line represents the linear best fit line using Pearson’s correlation. Blue shaded region denotes the variability of the line and the resulting estimates from Pearson’s correlation are shown in text above the shaded region.

**Figure S5:**
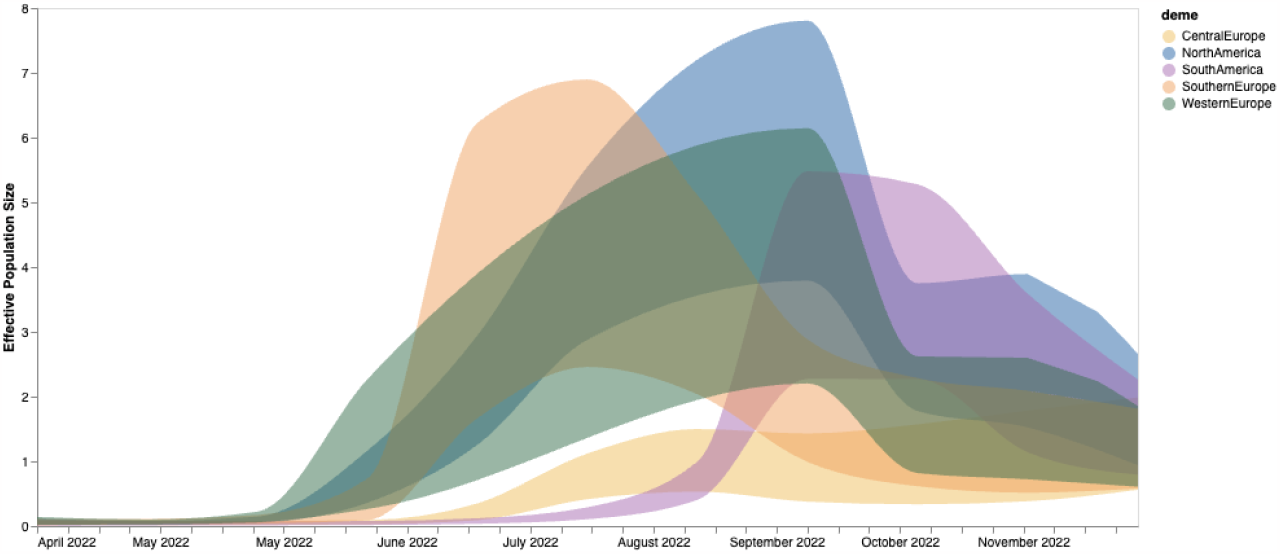
Effective population size estimated via MASCOT-Skyline, related to Figure 3. Estimates of effective population sizes (NeTao in years) from April 2022 through December 2024 using 587 sequences subsampled equally throughout time. In contrast to the main MASCOT-GLM analysis, no empirical predictors were used, showing the extent of phylogenetic signal and uncertainty when using only genomes.

**Figure S6:**
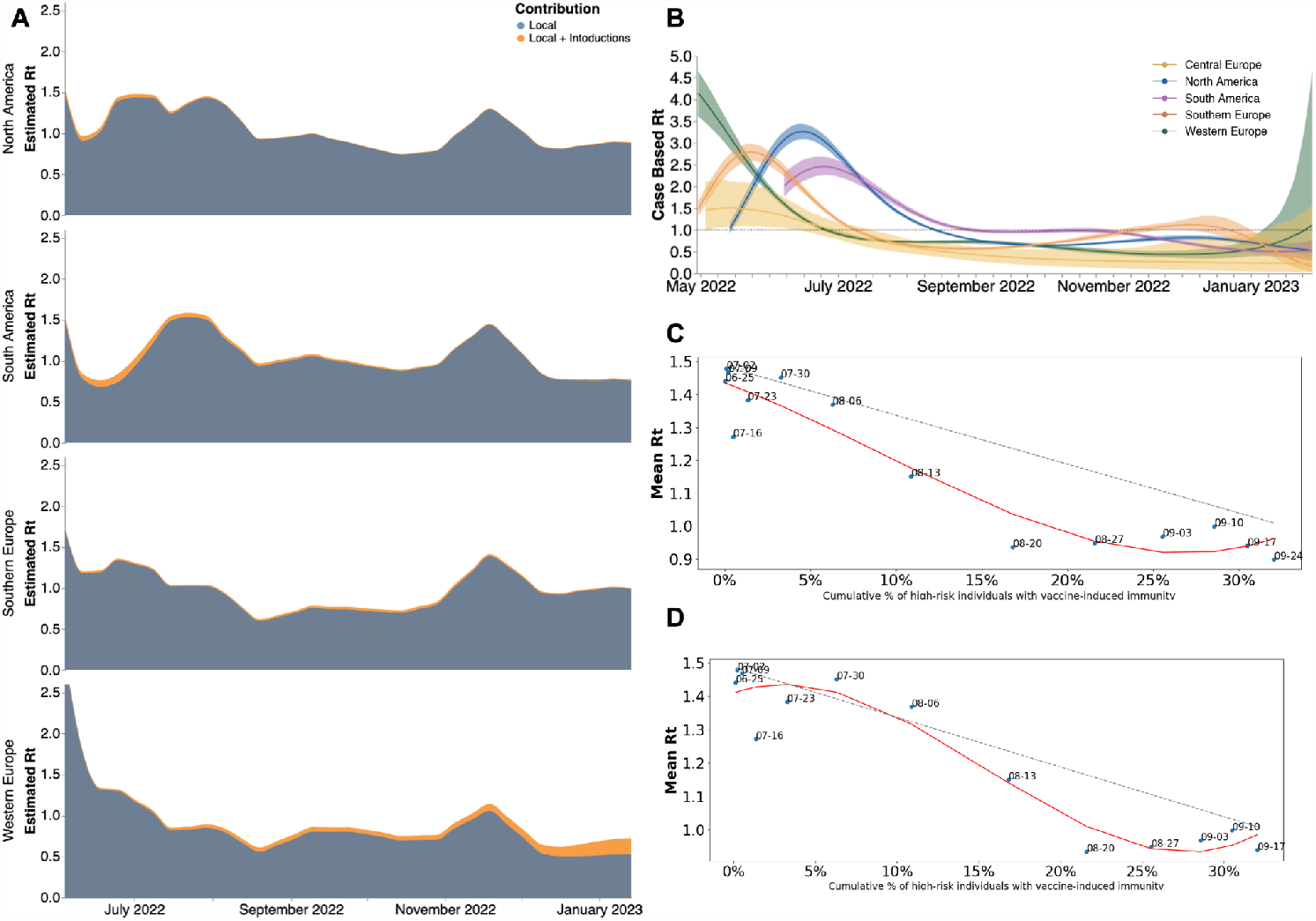
Estimates of time-varying reproductive number (Rt), related to figures 5 and 6. **(A)** Estimates of Rt from April 2022 through December 2023 via MASCOT-GLM for four global regions separated by source of contribution. Blue denotes local Rt without the influence of outside viral introductions while orange shows the added contribution of introductions. Central Europe was removed due to limited data on introductions. **(B)** Estimates of time-varying reproductive number (Rt) in five global regions Estimates of Rt from April 2022 through December 2022 using renewal model framework from case counts only. The inner area denotes the 50% HPD interval and the outer area denotes the 95% HPD interval. Dashed line highlights an Rt value of 1 above which denotes an exponentially growing viral epidemic. (**C-D**) Scatter plot comparing mean *Rt* calculated via MASCOT-GLM for North America vs cumulative percentage of high risk individuals with vaccine-induced immunity in the United States with a one week lag to account account for the development of immunity **(C)** and no lag following date of vaccination **(D)**. Red line indicates the best fit spline for scattered points. Dashed gray line indicates expected linear decrease in *Rt* with increasing vaccine-immunity assuming SIR dynamics. Over each point are the dates that correspond to the mean *Rt* and percent of immunity at that moment.

**Figure S7:**
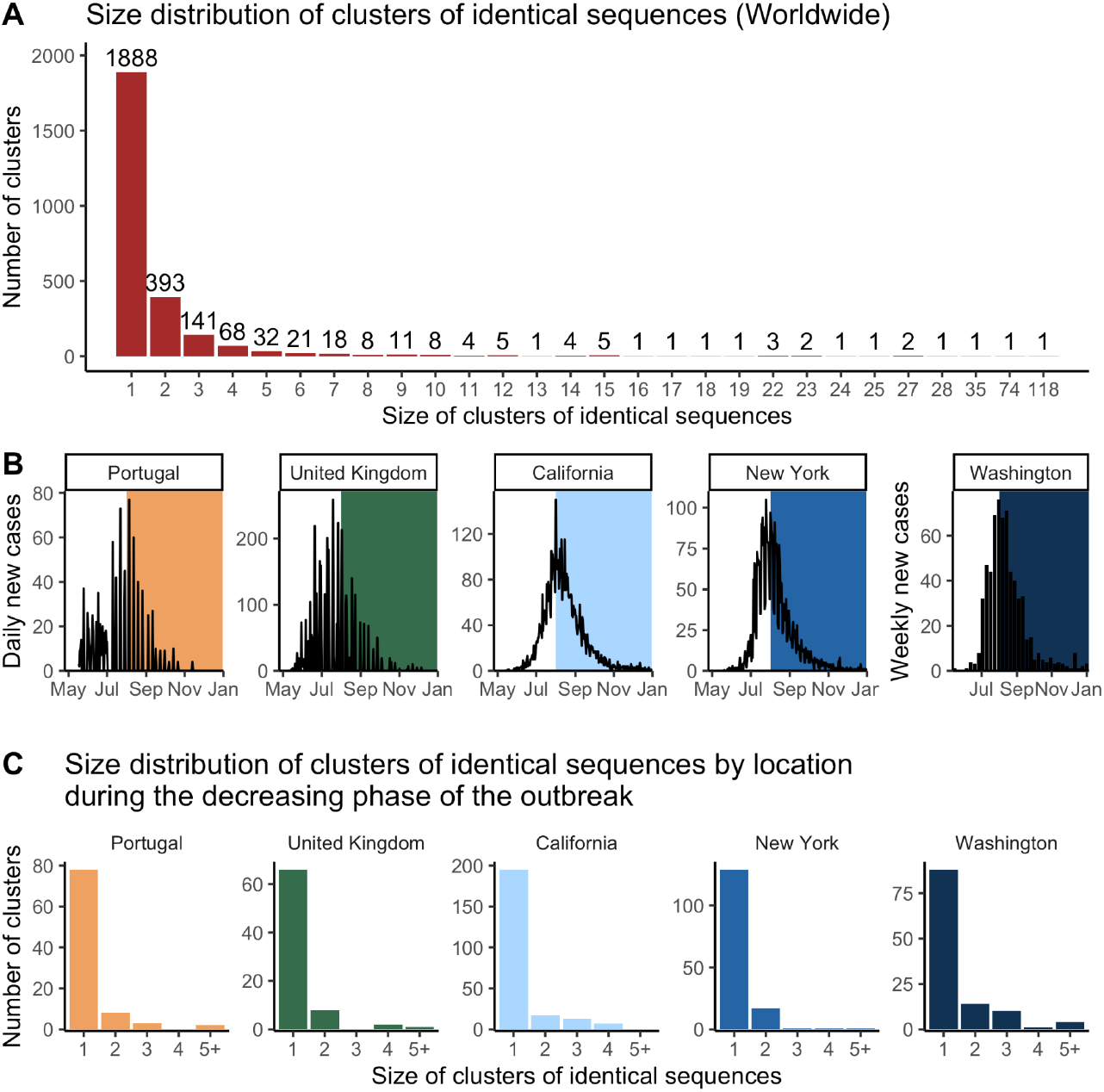
Size distribution of clusters of identical mpox sequences, related to Figure 7. **(A)** Size distribution of clusters of identical mpox sequences worldwide. **(B)** Dynamics of mpox cases in the location of study. The coloured rectangles correspond to the study period. **(C)** Size distribution of clusters of identical mpox sequences by location during the decreasing phase of the outbreak (study period defined in B).

**Table S1.** Geocoding for various country and regional scales used in this study, related to Figures 2 and 3. DTA denotes the samples for the phylogeographic analysis which was subsampled using confirmed case counts as weights. MASCOT-GLM column denotes the sample for the phylodynamic inference which was subsampled by enforcing equal temporal sampling per country per month.

| Region | Country | DTA | MASCOT-GLM |
| --- | --- | --- | --- |
| North America | Canada | 27 | 45 |
|  | United States | 533 | 120 |
| Western Europe | Austria | 3 | 11 |
|  | Germany | 116 | 84 |
|  | Switzerland | 7 | 7 |
|  | United Kingdom | 94 | 78 |
|  | France | 25 | 23 |
|  | Belgium | 15 | 18 |
| Central Europe | Slovakia | 0 | 11 |
|  | Slovenia | 0 | 31 |
| Southern Europe | Italy | 6 | 7 |
|  | Portugal | 88 | 75 |
|  | Spain | 9 | 11 |
| South America | Colombia | 77 | 57 |
|  | Peru | 3 | 10 |

**Table S2.**
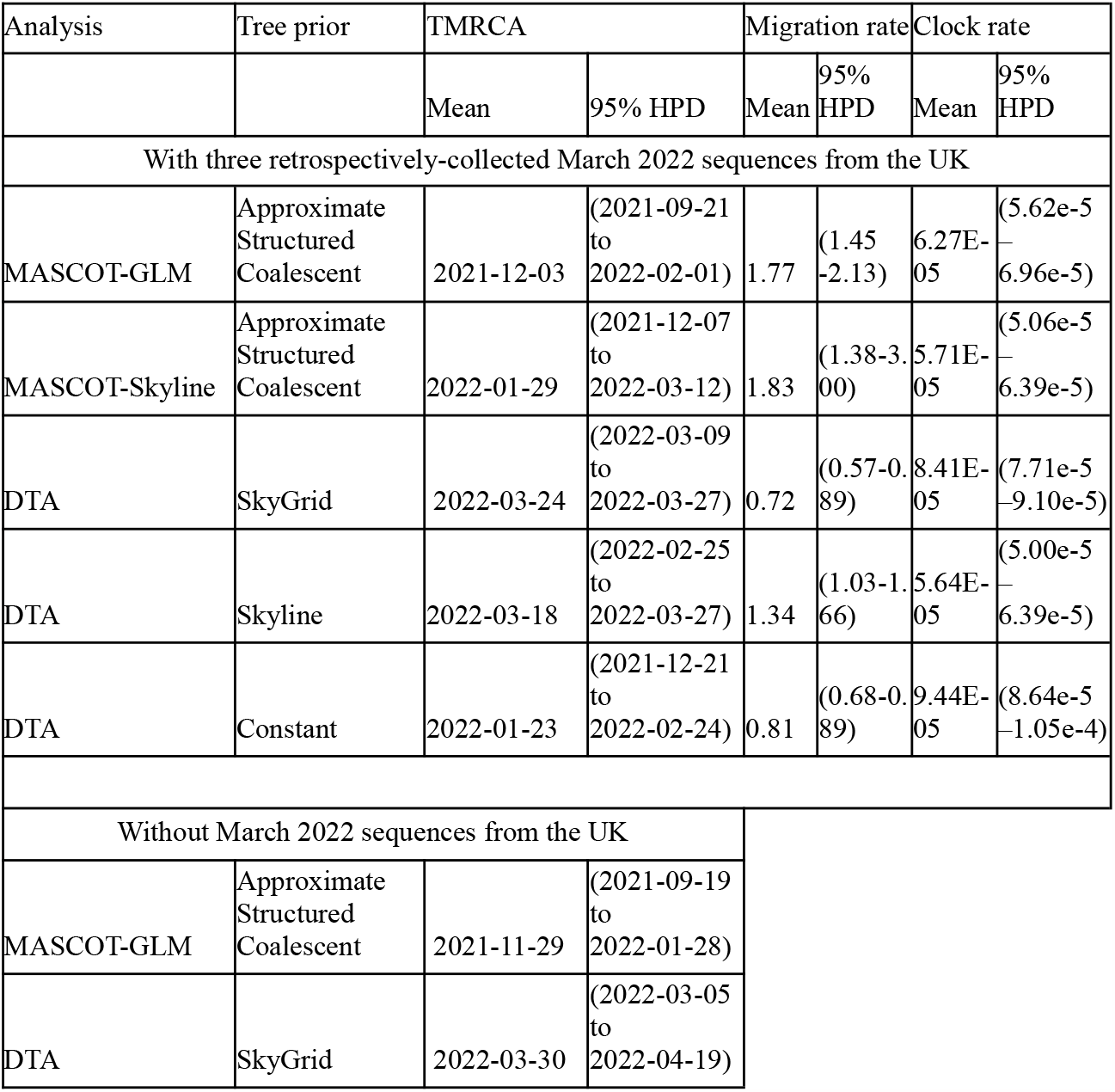
Comparison of time to most recent common ancestor (TMRCA), migration rate (migration events per year), and clock rate (substitutions per site per year) by method, related to Figures 2 and 3. First five rows denote the comparison of key summary statistics from main and alternative models used (with varying tree priors and inclusion of empirical predictors) which include three sequences from March 2022 which were found retrospectively in the UK. The last two rows represent the main analyses but without the three retrospective march 2022 samples.

| Analysis | Tree prior | TMRCA |  | Migration rate |  | Clock rate |  |
| --- | --- | --- | --- | --- | --- | --- | --- |
|  |  | Mean | 95% HPD | Mean | 95% HPD | Mean | 95% HPD |
| With three retrospectively-collected March 2022 sequences from the UK |  |  |  |  |  |  |  |
| MASCOT-GLM | Approximate Structured Coalescent | 2021-12-03 | (2021-09-21 to 2022-02-01) | 1.77 | (1.45-2.13) | 6.27E-05 | (5.62e-5 – 6.96e-5) |
| MASCOT-Skyline | Approximate Structured Coalescent | 2022-01-29 | (2021-12-07 to 2022-03-12) | 1.83 | (1.38-3.00) | 5.71E-05 | (5.06e-5 – 6.39e-5) |
| DTA | SkyGrid | 2022-03-24 | (2022-03-09 to 2022-03-27) | 0.72 | (0.57-0.89) | 8.41E-05 | (7.71e-5 – 9.10e-5) |
| DTA | Skyline | 2022-03-18 | (2022-02-25 to 2022-03-27) | 1.34 | (1.03-1.66) | 5.64E-05 | (5.00e-5 – 6.39e-5) |
| DTA | Constant | 2022-01-23 | (2021-12-21 to 2022-02-24) | 0.81 | (0.68-0.89) | 9.44E-05 | (8.64e-5 – 1.05e-4) |
| Without March 2022 sequences from the UK |  |  |  |  |  |  |  |
| MASCOT-GLM | Approximate Structured Coalescent | 2021-11-29 | (2021-09-19 to 2022-01-28) |  |  |  |  |
| DTA | SkyGrid | 2022-03-30 | (2022-03-05 to 2022-04-19) |  |  |  |  |

**Table S3.** Reproduction numbers and dispersion parameter estimates from the analysis of the size distribution of clusters of identical sequences using a joint-likelihood, related to Figure 7. For each location, we report maximum likelihood estimates (MLE) along 95% likelihood profile confidence intervals. Different assumptions regarding the proportion of infections sequenced are explored. These estimates were obtained by allowing the reproduction numbers to vary between regions but assuming a similar value of the dispersion parameter *k* across locations.

| Assumption regarding the proportion of infections detected | Location | $R$ estimate (maximum likelihood estimate with 95% confidence interval) | $k$ estimate (maximum likelihood estimate with 95% confidence interval) | Proportion of infected individuals with 0 offspring (obtained from $R$ and $k$ MLE) |
| --- | --- | --- | --- | --- |
| 10% | Portugal | 0.87 (0.71-1.03) | 0.16 (0.11-0.23) | 74% |
|  | United Kingdom | 1.10 (0.98-1.22) |  | 72% |
|  | California (USA) | 1.14 (1.08-1.20) |  | 72% |
|  | New York (USA) | 1.05 (0.94-1.14) |  | 72% |
|  | Washington (USA) | 1.06 (0.94-1.19) |  | 72% |
| 50% | Portugal | 0.60 (0.44-0.80) | 0.30 (0.18-0.54) | 72% |
|  | United Kingdom | 0.89 (0.73-1.06) |  | 66% |
|  | California (USA) | 0.94 (0.85-1.03) |  | 65% |
|  | New York (USA) | 0.82 (0.69-0.94) |  | 67% |
|  | Washington (USA) | 0.84 (0.69-1.02) |  | 67% |
| 100% | Portugal | 0.46 (0.31-0.66) | 0.36 (0.21-0.71) | 74% |
|  | United Kingdom | 0.76 (0.59-0.95) |  | 66% |
|  | California (USA) | 0.82 (0.72-0.92) |  | 65% |
|  | New York (USA) | 0.68 (0.55-0.82) |  | 68% |
|  | Washington (USA) | 0.70 (0.54-0.91) |  | 68% |

**Table S4.** Location-specific reproduction number and dispersion parameter estimates from the analysis of the size distribution of clusters of identical sequences, related to Figure 7. For each location, we report maximum likelihood estimates (MLE) along 95% likelihood profile confidence intervals. Different assumptions regarding the proportion of infections sequenced are explored.

| Location | Assumption regarding the proportion of infections detected | *R* estimate (maximum likelihood estimate with 95% confidence interval) | *k* estimate (maximum likelihood estimate with 95% confidence interval) | Proportion of infected individuals with 0 offspring (obtained from *R* and *k* MLE) |
| --- | --- | --- | --- | --- |
| Portugal | 10% | 0.8 (0.64-0.98) | 0.09 (0.02-0.3) | 81% |
|  | 50% | 0.57 (0.40-0.79) | 0.19 (0.04-1.8) | 77% |
|  | 100% | 0.45 (0.29-0.67) | 0.23 (0.06-6.5) | 78% |
| United Kingdom | 10% | 0.97 (0.81-1.12) | 0.11 (0.03-0.34) | 78% |
|  | 50% | 0.76 (0.58-0.95) | 0.23 (0.04-2.6) | 72% |
|  | 100% | 0.63 (0.45-0.84) | 0.28 (0.05- >10) | 72% |
| California (USA) | 10% | 1.03 (0.95-1.11) | 0.07 (0.03-0.11) | 83% |
|  | 50% | 0.89 (0.8-0.98) | 0.19 (0.08-0.42) | 72% |
|  | 100% | 0.79 (0.68-0.89) | 0.25 (0.1-0.75) | 70% |
| New York (USA) | 10% | 1.21 (1.14-1.28) | 1.26 (0.37 - >10) | 42% |
|  | 50% | 0.92 (0.79-1.03) | 1.04 (0.24- >10) | 52% |
|  | 100% | 0.75 (0.61-0.88) | 0.99 (0.19- >10) | 58% |
| Washington (USA) | 10% | 1.1 (0.99-1.21) | 0.23 (0.07-0.72) | 67% |
|  | 50% | 0.85 (0.71-1.02) | 0.38 (0.13-2.3) | 64% |
|  | 100% | 0.71 (0.55-0.9) | 0.43 (0.16-2.8) | 66% |

